# Efficacy and Cost of High-Frequency IGRT in Elderly Stage III Non-Small-Cell Lung Cancer Patients

**DOI:** 10.1101/2020.12.01.20241794

**Authors:** Samuel P Heilbroner, Eric P Xanthopoulos, Donna Buono, Daniel Carrier, Ben Y Durkee, Michael Corradetti, Tony JC Wang, Alfred I Neugut, Dawn L Hershman, Simon K Cheng

## Abstract

**Background:** High-frequency image-guided radiotherapy (hfIGRT) is ubiquitous but its benefits are unproven. We examined the cost effectiveness of hfIGRT in stage III non-small-cell lung cancer (NSCLC).

**Methods:** We selected stage III NSCLC patients ≥66 years old who received definitive radiation therapy from the Surveillance, Epidemiology, and End-Results-Medicare database. Patients were stratified by use of hfIGRT using Medicare claims. Predictors for hfIGRT were calculated using a logistic model. The impact of hfIGRT on lung toxicity free survival (LTFS), esophageal toxicity free survival (ETFS), cancer-specific survival (CSS), overall survival (OS), and cost of treatment was calculated using Cox regressions, propensity score matching, and bootstrap methods.

**Results:** Of the 4,430 patients in our cohort, 963 (22%) received hfIGRT and 3,468 (78%) did not. By 2011, 49% of patients were receiving hfIGRT. Predictors of hfIGRT use included treatment with intensity-modulated radiotherapy (IMRT) (OR = 7.5, p < 0.01), recent diagnosis (OR = 51 in 2011 versus 2006, p < 0.01), and residence in regions where the Medicare intermediary allowed IMRT (OR = 1.50, p < 0.01). hfIGRT had no impact on LTFS (HR 0.97; 95% CI 0.86 – 1.09), ETFS (HR 1.05; 95% CI 0.93–1.18), CSS (HR 0.94; 95% CI 0.84 – 1.04), or OS (HR 0.95; 95% CI 0.87 – 1.04). Mean radiotherapy and total medical costs six months after diagnosis were $17,330 versus $15,024 (p < 0.01) and $71,569 versus $69,693 (p = 0.49), respectively.

**Conclusion:** hfIGRT did not affect clinical outcomes in elderly patients with stage III NSCLC but did increase radiation cost.

## BACKGROUND

High-frequency radiation therapy (hfIGRT) is the process of imaging a patient before almost every treatment or fraction to improve the precision and accuracy of radiation delivery. If a patient’s tumor or vital organs become shifted from one treatment to another, imaging can identify and help correct this source of error. By reducing anatomic variation, hfIGRT has the potential to ensure adequate radiation dose to the tumor and reduce dose to the surrounding vital organs; thus increasing radiation efficacy and reducing toxicities.

Given the perceived benefits, hfIGRT is being rapidly adopted across the United States, particularly in lung cancer. A 2015 survey found that 78% of radiation oncologist use image guided radiation therapy (IGRT) on a daily basis in at least some of their lung cancer patients.^1^ But the evidence supporting the use of hfIGRT in lung cancer is limited. While studies have shown that IGRT reduces variation in patient position^2-11^ and computer simulations have shown that reducing that variation could impact clinical end points like tumor control^12-16^, there is little clinical evidence linking hfIGRT with better outcomes in lung cancer. Two small single institutional retrospective studies have shown that daily IGRT is associated with either improved survival and local control, though reported no toxicity results.^17,18^ More accurate treatments with hfIGRT also has the potential to decrease the margin size given for treatment setup uncertainty, the planning treatment volume (PTV), and thereby reduce toxicities.

The efficacy of hfIGRT in lung cancer is largely untested. Yet IGRT has been shown to increase cost.^1^ So what is the benefit of hfIGRT? There are no prospective randomized trials on IGRT technologies in lung cancers. Therefore, hfIGRT’s clinical efficacy and cost effectiveness are best examined using retrospective real world data. We used the Surveillance, Epidemiology, and End Results Database attached to Medicare (SEER-Medicare) to compare the efficacy (in terms of toxicity and survival) and cost of hfIGRT in stage III non-small cell lung cancer (NSCLC).

## METHODS

### Cohort Selection

We identified patients ≥66 years of age with pathologically confirmed stage III NSCLC diagnosed from January 1, 2006 to December 31, 2011 from the SEER-Medicare database. The SEER-Medicare database captures clinical and claims data for Medicare beneficiaries with cancer who reside within 16 geographic catchment areas representing about 35% of the US population. Patients were excluded if they had: (1) any prior malignancies; (2) autopsy or death certificate as the basis for the lung cancer diagnosis; (3) reason for Medicare entitlement other than age; (4) >90 days discordance between SEER and Medicare reported date of death; (5) any health maintenance organization enrollment within one year of diagnosis; (6) incomplete Part A or B Medicare insurance within one year of diagnosis; (7) or residence in a nursing home at the time of diagnosis. We only selected patients who (8) received definitive radiation treatment based on claims as described in the appendix **Section M1**. A consort diagram is provided in the appendix (**Table T1**).

### Definition of high-frequency image-guided radiotherapy (hfIGRT)

We subsequently divided our cohort into patients who received hfIGRT and those who did not. Imaging was identified by looking for claim codes (appendix **Table T2**) for portal X-rays, orthogonal X-rays, or cone beam computed tomography (CBCT) imaging from the 15 days prior to diagnosis through the 15 days after the end of radiotherapy. Since CBCT is the most common form of IGRT in practice, we also restricted our definition of IGRT to CBCT alone as a sensitivity analysis. Results were nearly identically to those below with no statistical difference. hfIGRT was defined as imaging with ≥ 65% of radiation fractions. This cutoff is validated in the appendix (**Section M2, Figure F1**).

### Other Covariates

Using SEER-registry and claims data we also analyzed age at diagnosis, sex, race, marital status, state, year of diagnosis, the Charlson comorbidity index, medical history of COPD, use of supplemental oxygen, homebound status, primary tumor characteristics (e.g. histology, stage, size, and location), treatment using intensity-modulated radiation therapy (IMRT), the number of radiation fractions delivered, whether the patient received therapy in a hospital or freestanding clinic, whether the patient received care in a rural or urban setting, type of treatment (e.g. chemoradiation, trimodality therapy, surgery and radiation, or radiation alone), and the total volume of patients seen by a patient’s radiation oncologist. A detailed description of these covariates and how they were calculated is in the appendix **Section M3** and uses several references.^19-26^

### Efficacy outcomes

The primary end points were overall survival, cancer-specific survival, lung cancer-specific survival, acute esophageal toxicity rates, and acute pulmonary toxicity rates. As described in Shirvani et al^27^, we assessed lung toxicity using a “narrow” definition included only the ICD-9 diagnosis code for “unspecified acute pulmonary toxicity due to radiation” and a “broad” definition that also included non-radiation-related toxicity codes (i.e. pneumonia). For acute esophagitis, we evaluated codes for (1) esophagitis, (2) dehydration, (3) feeding tube placement, and (4) mucositis, individually and in aggregate. Toxicity codes are listed in the appendix **Table T4**.

### Cost of hfIGRT

Adopting the method pioneered by Smith et al,^28^ we calculated the total cost of care and the cost of radiotherapy by summing the total amount reimbursed by Medicare to the provider over the interval from 15 days prior to diagnosis to 6 months after diagnosis. See the appendix **Section M4** for details.

### Effect of Local Coverage Determination on hfIGRT use and Cost

To determine if governmental insurance policies limited the use and cost of hfIGRT, we used Medicare carrier local coverage determinations (LCDs). LCDs are regional policy guidelines that describe when a service is covered. There are no LCDs governing IGRT use. However, in practice IMRT, another advanced radiation technology, and hfIGRT are tightly correlated. Hence, we used LCDs for IMRT as a proxy for hfIGRT, binning them into LCDs restrictive of IMRT, LCDs permissive of IMRT, and LCDs without IMRT guidance.^28^ Details are in the appendix **Section M5**.

### Statistical Analyses

Statistical methods are described in detail in the appendix **Section M6**. Briefly, we compared the distribution of patient characteristics between the two treatment groups with the Pearson’s chi-square test. Predictors of hfIGRT utilization were calculated using two logistic models that used two different proxies for geographic location: state and LCD. The association between hfIGRT and clinical outcomes (e.g. overall survival, cancer-specific survival, and toxicities) was assessed using Kaplan-Meier curves and multivariate Cox modeling accounting for the covariates described above.

Based on the results of our state-based logistic regression, we created a propensity score model to validate the findings of the multivariate Cox regressions. hfIGRT patients were matched 1-to-1 without replacement with non-hfIGRT counterparts. Proportional hazards models were used to adjust for covariates that could not be balanced using matching. Kaplan-Meier plots for the matched cohorts were also generated.

A non-parametric bootstrap model was used to estimate the 95% confidence interval (CI) around the mean cost difference between the hfIGRT and non-hfIGRT groups. We estimated these differences for both the original cohort and the 1-to-1 matched sub-cohort. The impact of individual covariates on the cost of treatment was assessed using the Wilcoxon two-sample test. Because costs were calculated during the 6 months after diagnosis, only patients who lived at least 6 months from diagnosis were included in the cost analysis. The 1-to-1 matched sub-cohort used in the cost analysis was generated from a logistic regression that only included these 6-month survivors.

## RESULTS

### Patient characteristics

4,430 patients satisfied the inclusion criteria for our study. Of these, 963 (22%) patients received hfIGRT and 3,468 (78%) patients did not (appendix **Table T1**).

### Trends in Utilization of hfIGRT

Billing for hfIGRT increased from 2% of patients diagnosed in 2006 to 49% in 2011. Regions with permissive IMRT LCDs did adopt hfIGRT significantly faster than restrictive LCDs (p < 0.01, **Figure 1a**). In contrast, many other factors, like the type of healthcare center (freestanding clinic versus hospital based), were not associated with the adoption of hfIGRT (p = 0.18, **Figure 1b**).

**Figure 1:**
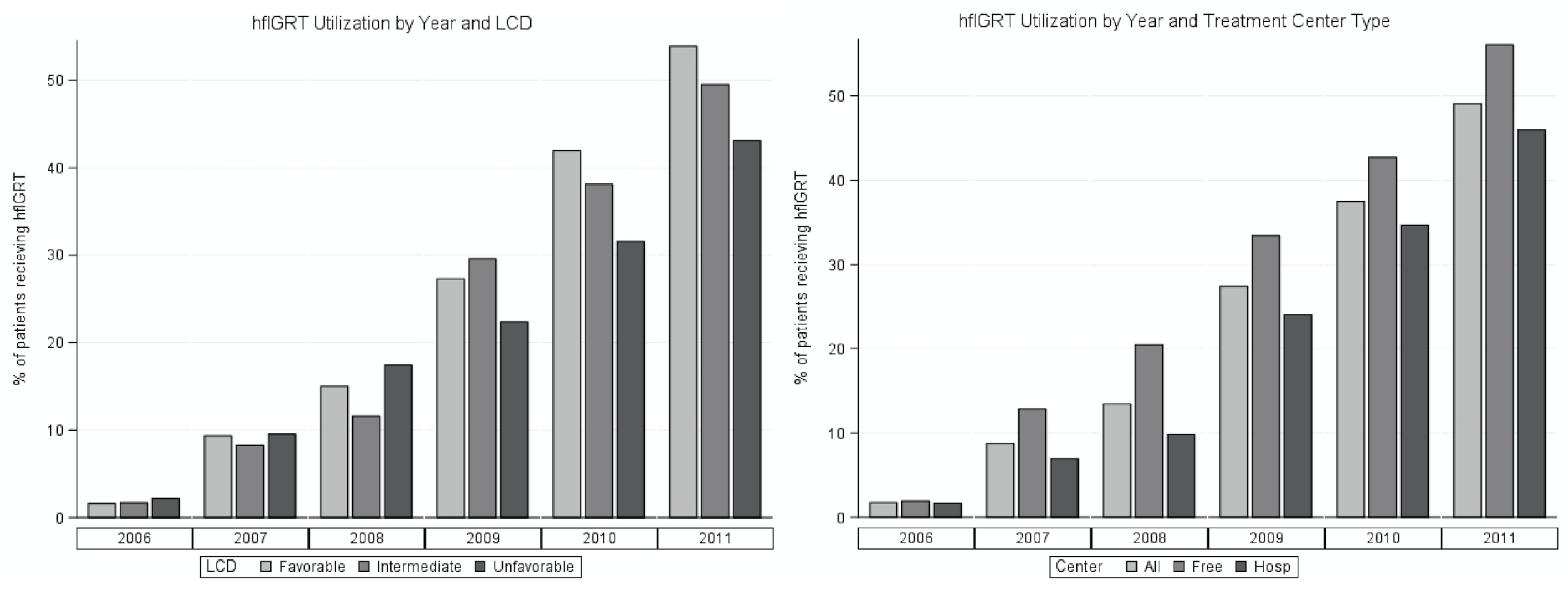
(A) Percent of stage IIIA NSCLC patients receiving hfIGRT in free-standing and hospital based practices by year. There was a trend toward increased utilization among free-standing radiation oncology facilities, but this did not reach significance (Figure 1a). (B) Percent of stage IIIA NSCLC who received hfIGRT in regions with permissive LCDs, restrictive LCDs, or no LCDs governing IMRT use. Although IMRT is distinct of hfIGRT, the two are highly correlated. Restrictive LCDs were associated with a significant reduction if hfIGRT use (p < 0.01). Abbreviations: NSCLC = non-small cell lung cancer, hfIGRT = high frequency image guided radiation therapy, LCD = local coverage determination, IMRT = intensity-modulated radiation therapy.

CBCT rapidly overtook portal and KV X-ray films as the imaging modality of choice for hfIGRT (appendix **Figure F2**). Of patients who received hfIGRT, most (≥60%) received CBCT imaging with every radiation treatment (appendix **Figure F3**).

In our study, most physicians used hfIGRT in a binary fashion: for either all of their patients or for none of them. Specifically, 73% of physicians never used hfIGRT in any of their patients. Of the 27% who did use hfIGRT, the most common practice was to use hfIGRT in all (100%) of their stage III NSCLC patients (appendix **Figure F4**).

### Multivariable logistic model predicting odds ratios (OR) of hfIGRT use

A state-based multivariable logistic model identified multiple associations with hfIGRT (**Table 1**). hfIGRT was very strongly associated with IMRT (OR = 7.51, p <0.01). hfIGRT billing also increased significantly with the year of diagnosis (OR = 51 for 2011 versus 2006, p <0.01). hfIGRT use decreased with increased radiation oncologist density (OR = 0.57 in the 4^th^ quartile, p < 0.01); but it increased with increased general surgeon density (OR = 2.44 in the 4^th^ quartile, p < 0.01). A logistic regression using LCD as a proxy for location instead of state showed that hfIGRT was also rarer in jurisdictions with unfavorable LCDs for IMRT (OR = 0.66, p < 0.01, appendix **Table T4**).

**Table 1.**
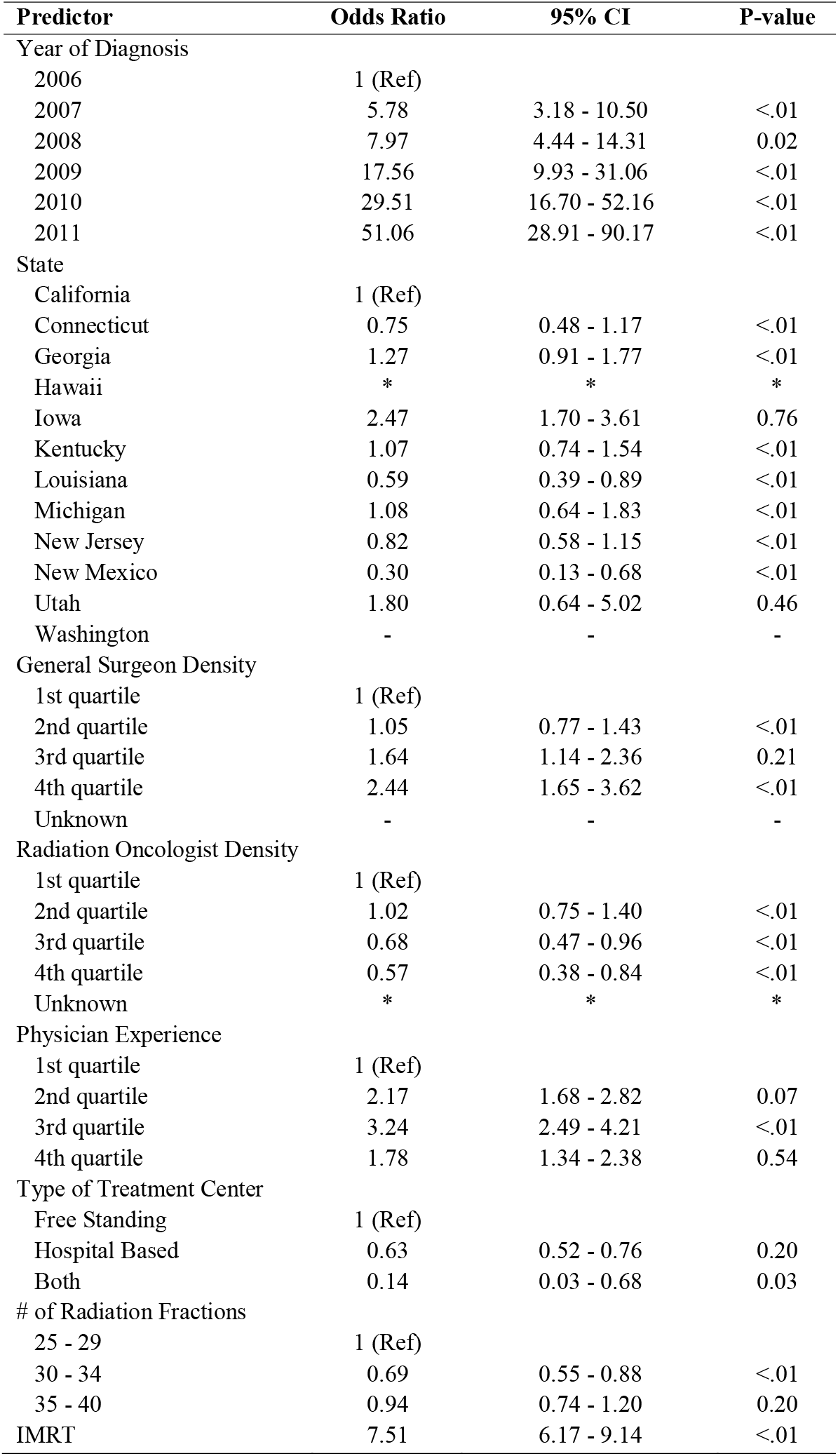
Logistic Regression Using State for Location. Predictors of billing for high frequency image guided radiation therapy (hfIGRT): multivariable model.

| **Predictor** | **Odds Ratio** | **95% CI** | **P-value** |
| --- | --- | --- | --- |
| Year of Diagnosis |  |  |  |
| 2006 | 1 (Ref) |  |  |
| 2007 | 5.78 | 3.18 - 10.50 | <.01 |
| 2008 | 7.97 | 4.44 - 14.31 | 0.02 |
| 2009 | 17.56 | 9.93 - 31.06 | <.01 |
| 2010 | 29.51 | 16.70 - 52.16 | <.01 |
| 2011 | 51.06 | 28.91 - 90.17 | <.01 |
| State |  |  |  |
| California | 1 (Ref) |  |  |
| Connecticut | 0.75 | 0.48 - 1.17 | <.01 |
| Georgia | 1.27 | 0.91 - 1.77 | <.01 |
| Hawaii | * | * | * |
| Iowa | 2.47 | 1.70 - 3.61 | 0.76 |
| Kentucky | 1.07 | 0.74 - 1.54 | <.01 |
| Louisiana | 0.59 | 0.39 - 0.89 | <.01 |
| Michigan | 1.08 | 0.64 - 1.83 | <.01 |
| New Jersey | 0.82 | 0.58 - 1.15 | <.01 |
| New Mexico | 0.30 | 0.13 - 0.68 | <.01 |
| Utah | 1.80 | 0.64 - 5.02 | 0.46 |
| Washington | - | - | - |
| General Surgeon Density |  |  |  |
| 1st quartile | 1 (Ref) |  |  |
| 2nd quartile | 1.05 | 0.77 - 1.43 | <.01 |
| 3rd quartile | 1.64 | 1.14 - 2.36 | 0.21 |
| 4th quartile | 2.44 | 1.65 - 3.62 | <.01 |
| Unknown | - | - | - |
| Radiation Oncologist Density |  |  |  |
| 1st quartile | 1 (Ref) |  |  |
| 2nd quartile | 1.02 | 0.75 - 1.40 | <.01 |
| 3rd quartile | 0.68 | 0.47 - 0.96 | <.01 |
| 4th quartile | 0.57 | 0.38 - 0.84 | <.01 |
| Unknown | * | * | * |
| Physician Experience |  |  |  |
| 1st quartile | 1 (Ref) |  |  |
| 2nd quartile | 2.17 | 1.68 - 2.82 | 0.07 |
| 3rd quartile | 3.24 | 2.49 - 4.21 | <.01 |
| 4th quartile | 1.78 | 1.34 - 2.38 | 0.54 |
| Type of Treatment Center |  |  |  |
| Free Standing | 1 (Ref) |  |  |
| Hospital Based | 0.63 | 0.52 - 0.76 | 0.20 |
| Both | 0.14 | 0.03 - 0.68 | 0.03 |
| # of Radiation Fractions |  |  |  |
| 25 - 29 | 1 (Ref) |  |  |
| 30 - 34 | 0.69 | 0.55 - 0.88 | <.01 |
| 35 - 40 | 0.94 | 0.74 - 1.20 | 0.20 |
| IMRT | 7.51 | 6.17 - 9.14 | <.01 |

### Cox proportional hazards regression analysis of survival and acute toxicities

On Cox regression analysis, hfIGRT was not associated with overall survival (HR 0.95; 95% CI 0.87 – 1.04), cancer-specific survival (HR 0.94; 95% CI 0.84 – 1.04), or lung toxicity-free survival under either our “narrow” (HR 0.81, 95% CI 0.62 – 1.05) or “broad” (HR 0.97, 95% CI 0.86–1.09) pulmonary toxicity definitions (**Table 2** and **Figure 2**).

**Table 2.** Association of hfIGRT with toxicities and survival, grouped by analytic technique.

| Outcome | HR | 95% CI | P-value |
| --- | --- | --- | --- |
| **Multivariate proportional hazards analysis<sup>a</sup>** |  |  |  |
| Overall survival | 0.95 | 0.87 - 1.04 | 0.29 |
| Cancer Specific Survival | 0.94 | 0.84 - 1.04 | 0.24 |
| Lung toxicity (Narrow) | 0.81 | 0.62 - 1.05 | 0.11 |
| Lung toxicity (Broad) | 0.97 | 0.86 - 1.09 | 0.60 |
| Esophageal toxicity (All) | 1.05 | 0.93 - 1.18 | 0.44 |
| Esophagitis/dysphagia | 1.27 | 1.08 - 1.49 | <.01 |
| Dehydration | 1.06 | 0.92 - 1.21 | 0.41 |
| Feeding tube placement | 1.07 | 0.74 - 1.54 | 0.72 |
| Mucositis | 1.70 | 1.03 - 2.80 | 0.04 |
| **Propensity score matching analysis<sup>b</sup>** |  |  |  |
| Overall survival | 0.95 | 0.85 - 1.07 | 0.42 |
| Cancer specific survival | 0.93 | 0.82 - 1.07 | 0.32 |
| Lung toxicity (Narrow) | 0.79 | 0.56 - 1.12 | 0.19 |
| Lung toxicity (Broad) | 1.02 | 0.87 - 1.21 | 0.78 |
| Esophageal toxicity (All) | 1.06 | 0.91 - 1.24 | 0.46 |
| Esophagitis/dysphagia | 1.21 | 0.97 - 1.51 | 0.09 |
| Dehydration | 1.06 | 0.89 - 1.27 | 0.52 |
| Feeding tube placement | 1.28 | 0.74 - 2.21 | 0.38 |
| Mucositis | 1.36 | 0.68 - 2.72 | 0.38 |

<sup>a</sup> Hazard ratios measure the effect of IGRT compared to control (referent) for each outcome
listed after adjustment for pre-specified clinically relevant covariates and covariates significant at $P < 0.20$ in univariate analysis. A separate model was created for each of the seven outcomes listed (Supplementary Tables T5–T13).
<sup>b</sup> Hazard ratios measure the effect of IGRT compared to control (referent) using proportional hazards regression of propensity-score matched hfIGRT cases and controls. Unmatched covariates were adjusted for using multivariate proportional hazard models.

**Figure 2:**
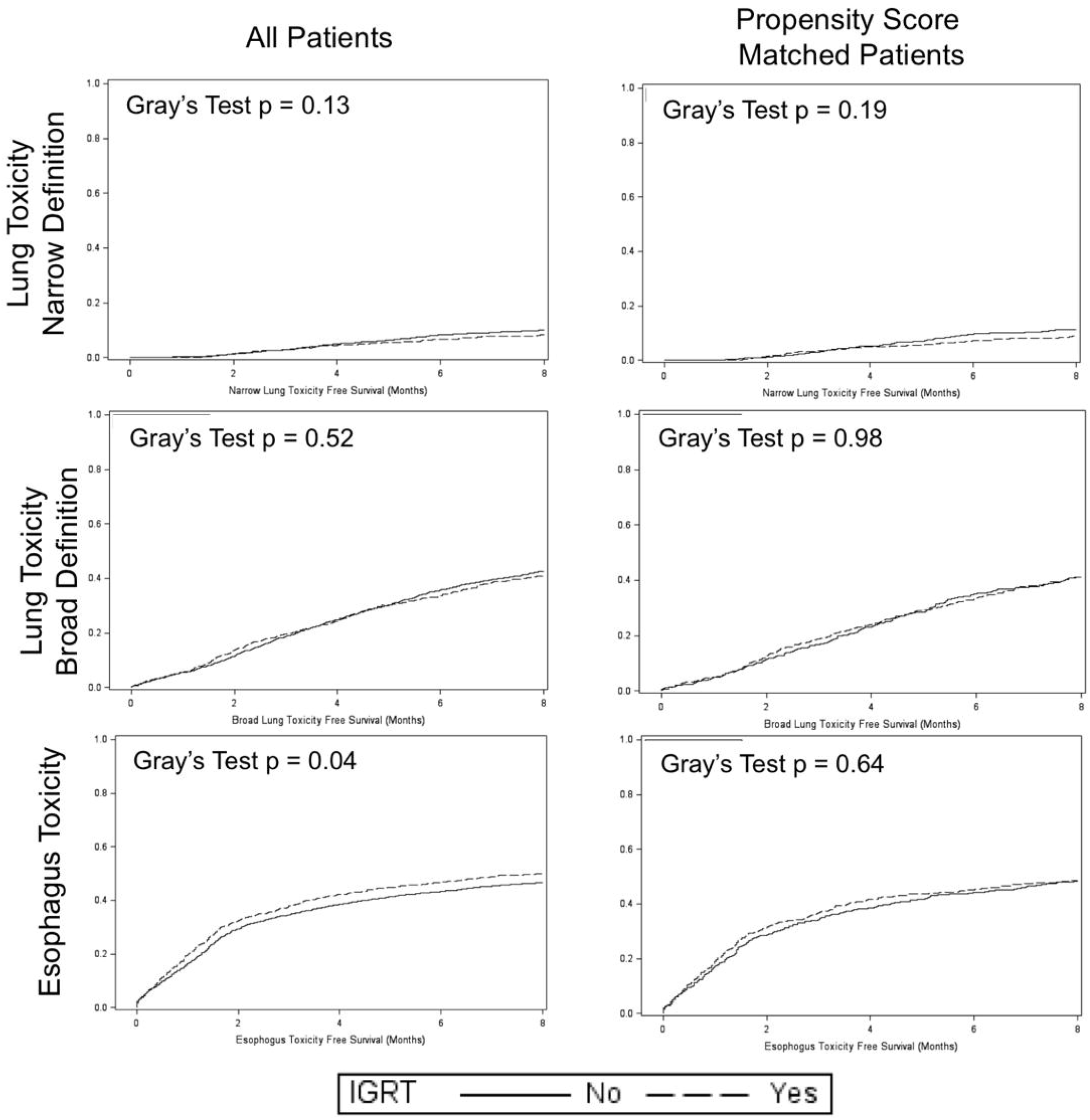
Cumulative incidence of acute toxicities calculated via claims for hfIGRT (dashed line) and other (solid line). No statistically significant difference was observed for any of the endpoints except esophagus toxicity. Although hfIGRT was associated with increased esophageal toxicity among all patients, this association did not withstand and analysis using propensity score matching. Abbreviations: hfIGRT = high frequency image guided radiation therapy.

hfIGRT was associated with increased risk of esophagitis alone (HR 1.27, 95% CI 1.08 – 1.49) and mucositis (HR 1.70, 95% CI 1.03 – 2.80). But - as described in the next section – these associations faded on propensity score-matched sub-analysis. hfIGRT was not associated with dehydration (HR 1.06, 95% CI 0.92 – 1.21), feeding tube placement (HR 1.07, 95% CI 0.74 – 1.54), or aggregated esophageal toxicity (HR 1.05, 95% CI 0.93 – 1.18).

Many other covariates besides hfIGRT were associated with toxicity and survival (appendix **Tables T5 – T13**). For example, IMRT use associated with aggregated esophageal toxicity (HR 1.20, 95% CI 1.06 – 1.34) (appendix **Table T7**); dehydration (HR 1.18, 95% CI 1.03 – 1.35) (appendix **Table T9**); reduced cancer-specific survival (HR 1.13, 95% CI 1.03 – 1.24) (appendix **Table T12**); and reduced overall survival (HR 1.12, 95% CI 1.03 – 1.22) (appendix **Table T13**).

### Propensity score-matched analysis of survival and acute toxicities

713 (74%) of the hfIGRT patients were successfully matched via propensity scoring to a control. The paired cohorts were well-balanced (p > 0.20) across all covariates except race (Table T3). Repeat multivariable Cox regressions on our matched cohort found no association between hfIGRT and any clinical endpoint, including overall (HR 0.95, 95% CI 0.85 - 1.07) or cancer-specific (HR 0.93, 95% CI 0.82 - 1.07) survival; “narrow” (HR 0.79, 95% CI 0.56 - 1.12) or “broad” (HR 1.02, 95% CI 0.87 - 1.21) lung toxicity-free survival; or esophagitis alone (HR 1.21, 95% CI 0.97 – 1.51), dehydration (HR 1.06, 95% CI 0.89 – 1.27), feeding tube placement (HR 1.28, 95% CI 0.74 – 2.21), mucositis (HR 1.36, 95% CI 0.68 – 2.72), or aggregated esophageal toxicity (HR 1.06, 95% CI 0.91 – 1.24) (Table 2). Cumulative toxicity hazard functions for the matched and unmatched cohorts are shown in **Figure 2**. Results are similar to the Cox regressions. Kaplan-Meier curves for OS and CSS are not shown, but results are similar to the Cox regressions described above.

### Cost of therapy

Of our original 4,430 patients, 3,849 patients – including 852 hfIGRT and 2,997 non-hfIGRT patients - lived at least six months and could be included in a cost analysis. This “cost-intended” cohort was re-matched 1:1 as described above. Covariates were well balanced across all covariates except race and sex (appendix **Table T14**). However, in the case of race and sex, the absolute difference the bivariate distributions were small (< 6% for all).

In the cost-intended propensity-matched patient cohort, the mean total cost of radiation therapy within the first six months of diagnosis was $17,330 in hfIGRT patients and $15,024 in non-hfIGRT patients (difference = $2,306, 95% CI $1,559 - $3,047, p <0.01) (appendix **Table T15**). The mean total cost of medical care over that time was $71,569 in hfIGRT patients and $69,693 in non-hfIGRT patients (difference = $1,875, 95% CI -$3,231 - $6,925, p = 0.49). As an exploratory analysis, we also examined the cost of a patient’s care that was directly attributable to IGRT by simply adding up all the bills for image-guidance. The mean total cost of image guidance was $1,971 in hfIGRT patients and $199 in non-hfIGRT patients (difference = $1,772, 95% CI $1,606 - $1,926, p <0.01). From 2006 – 2011, total radiotherapy costs increased by about 40% in hfIGRT patients versus about 35% in non-hfIGRT patients (**Figure 3**). There was a statistically significant increase in mean IGRT costs in IMRT favorable ($714) versus restrictive ($611) LCDs (p < 0.01 with Wilcoxon test; appendix **Figure F5**).

**Figure 3:**
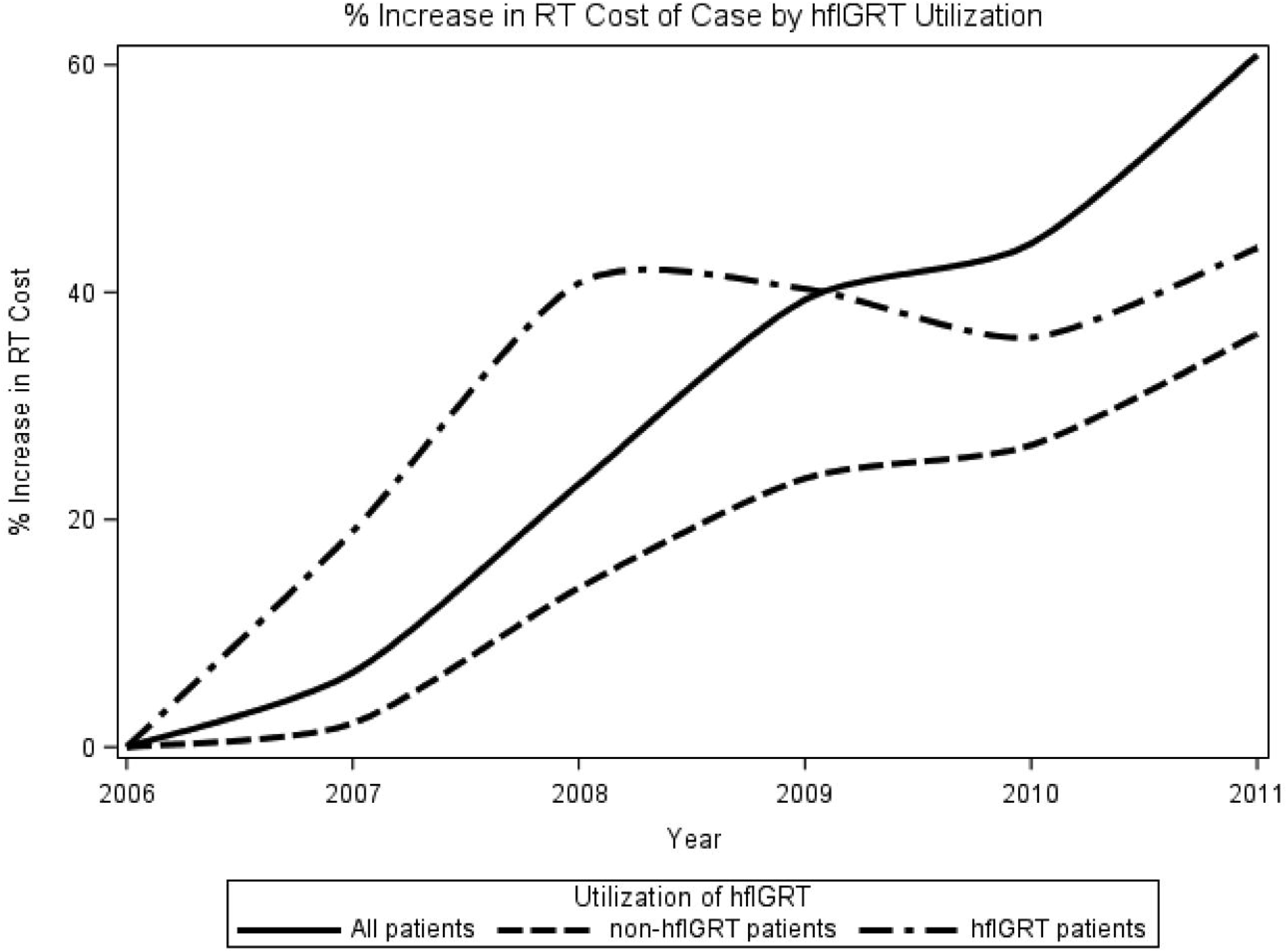
Percentage increase in mean radiation-related treatment cost. Costs include all Medicare reimbursements to providers for radiation therapy-related claims from 15 days before diagnosis to 6 months after diagnosis, are normalized to 2017 dollars, and are adjusted for geographic variation in Medicare reimbursement. Abbreviations: hfIGRT = high frequency image guided radiation therapy.

## DISCUSSION

Our study found a rapid increase in the utilization of hfIGRT among elderly Medicare patients with Stage III NSCLC; from just 2% in 2006 to 49% in 2011. Despite increased use, there has not been an increase in benefit. hfIGRT was not associated with improved survival or reduced rates toxicity. But hfIGRT was associated with an additional $2,306 in radiation related cost.

Costly advanced radiation technologies are often not subjected to rigorous scientific investigations for safety, efficacy or cost-effectiveness before wide-spread adoption into radiation oncology practice.^29,30^ Frequent IGRT has the potential to enhance the accuracy and precision of radiation delivery, however it is unclear if this translates to improved outcomes or lower costs. Two recent randomized controlled studies of hfIGRT for prostate cancer have shown mixed results. In a Norwegian prostate cancer study, daily IGRT was not associated with improved gastrointestinal or urinary toxicities nor survival outcomes; while in French study, daily IGRT improved rectal toxicity but had no statistically significant difference in the primary endpoint of recurrence-free survival. ^31,32^ Our study which is the largest analyzing the effects of hfIGRT on clinical outcomes in locally advanced lung cancer does not support improved survival nor decreased toxicity.

It is possible that patient selection bias could explain our results: Physicians may only prescribe IGRT to treat patients at the highest risk of toxicity. However, using Cox regression and matching analysis, we accounted for numerous clinical covariates that may impact toxicity and survival. Furthermore, our data suggests that most doctors are not prescribing IGRT on a case-by-case basis in a manner that would cause selection bias. Instead, the majority of physicians either consistently prescribe IGRT in all of their patients or never do.

One possible reason for hfIGRT’s failure to reduce toxicity in our study may be that in this early adoption period radiation oncologists did not reduce PTV margins despite using more frequent IGRT. Indeed, some recent reports suggest that radiation oncologists do not always pair more frequent IGRT use with PTV margin reductions and – as a result – may not always see clinical benefit from higher frequency IGRT use.^1^ As our dataset does not include PTV margin size, we are unable to analyze this possibility.

While IGRT did not affect outcomes, we did find that hfIGRT increased treatment imaging costs from about $200 in non-hfIGRT patients to about $2,000 in hfIGRT patients. Similarly, hfIGRT increased total radiation costs by about $2,000 in the 6-month interval following diagnosis, an increase of about 13%. Mean medical costs also increased by about $2000, but this difference did not achieve significance. We attribute this lack of significant to the higher variance associated with total medical costs.

Adoption of IGRT billing codes appears to have accelerated growth in the cost of radiotherapy for three reasons. First, as physicians embrace IGRT, they simply use it more: the number of IGRT fractions increased by nearly 700%, to 19,388 IGRT fractions in 2011, up from just 2,833 IGRT fractions in 2006 (appendix **Figure F2**). Second, as physicians are increasingly using more expensive forms of IGRT. Oncologists can perform IGRT with relatively inexpensive planar imaging or substantially more expensive (CBCT) volumetric imaging. Just 8% of IGRT fractions were CBCTs in 2006, compared to 86% in 2011. For reference, Varian pegged global Medicare reimbursements at $77 per planar image and $120 per volumetric image in 2017. Third, permissive (IMRT) LCDs appear to inflate both the rate and unit cost of IGRT use. Permissive LCDs are 1.5 times more likely to use hfIGRT (appendix **Table T4**) and permissive LCDs have per patient IGRT costs that are 17% higher than their restrictive counterparts (**Figure F5**). Taken together, lung cancer IGRT grew simultaneously more popular and more expensive from 2006 to 2011.

Reimbursement policy might help control costs. Although there are no LCDs governing IGRT use, we found that IMRT and IGRT billing are highly correlated. This enabled us to use LCDs covering IMRT as a proxy for IGRT, and these LCDs did significantly impact total billing per capita for IGRT. LCDs governing IGRT use may be even more effective.

Our study has several limitations. First, it included only elderly patients who are Medicare beneficiaries; the possibility remains that younger patients may not receive hfIGRT due to insurance coverage issues or amplified concerns of late toxic effects. Second, our definition of radiotherapy fraction hinged on the unvalidated method of counting radiotherapy claims for fraction delivery; but, Medicare claims are believed to be accurate because they are tied to physician payment. Third, our cost findings may not accurately reflect the cost of radiotherapy for patients with private insurance.

In conclusion, this study suggests that high-frequency IGRT increases radiotherapy costs without a commensurate improvement in clinical outcomes. Stricter treatment guidelines that encourage a reduction in PTV margins may help hfIGRT reach its full potential.

## Data Availability

This study makes use of the SEER-Medicare database, which is publically available through the NIH.

## ACKNOWLEDGMENTS

We thank Dr Tom K Hei, PhD, Vice-Chair and Professor of Radiation Oncology and Environmental Health Sciences, Columbia University for comments that greatly improved the manuscript. The authors acknowledge the efforts of the National Cancer Institute, the Office of Research, Development, and Information, Centers for Medicare & Medicaid Services; Information Management Services, Inc.; and the SEER program tumor registries in the creation of the SEER-Medicare database. Finally, the authors would like to endorse the following potential conflicts of interest: Dr. Wang reports personal fees and non-financial support from AbbVie, non-financial support from Merck, personal fees from AstraZeneca, personal fees from Doximity, personal fees and non-financial support from Novocure, personal fees and non-financial support from Elekta and personal fees from Wolters Kluwer, outside the submitted work; Dr. Cheng reports consulting fees from AbbVie. The other authors have no conflicts of interest.

## Abbreviations

CBCT: Cone-beam Computer Tomography
CI: Confidence Interval
CSS: Cancer-specific Survival
ETFS: Esophageal Toxicity Free Survival
hfIGRT: High-frequency Image Guided Radiation Therapy
IMRT: Intensity-modulated Radiotherapy
LCDs: Local Coverage Determinations
LTFS: Lung Toxicity Free Survival
NSCLC: Non-small-cell Lung Cancer
OR: Odds Ratios
OS: Overall Survival
SEER-Medicare: Surveillance, Epidemiology, And End-Results-Medicare

## REFRENCES

1. Nabavizadeh N, Elliott DA, Chen Y, et al. Image guided radiation therapy (IGRT) practice patterns and IGRT’s impact on workflow and treatment planning: Results from a national survey of American Society for Radiation Oncology members. International Journal of Radiation Oncology* Biology* Physics. 2016;94(4):850–857.

2. Bissonnette J-P, Purdie TG, Higgins JA, Li W, Bezjak A. Cone-beam computed tomographic image guidance for lung cancer radiation therapy. International Journal of Radiation Oncology* Biology* Physics. 2009;73(3):927–934.

3. Li W, Moseley DJ, Bissonnette J-P, Purdie TG, Bezjak A, Jaffray DA. Setup reproducibility for thoracic and upper gastrointestinal radiation therapy: Influence of immobilization method and on-line cone-beam CT guidance. Medical Dosimetry. 2011;35(4):287–296.

4. Chung PW, Haycocks T, Brown T, et al. On-line aSi portal imaging of implanted fiducial markers for the reduction of interfraction error during conformal radiotherapy of prostate carcinoma. International Journal of Radiation Oncology* Biology* Physics. 2004;60(1):329–334.

5. Zeidan OA, Langen KM, Meeks SL, et al. Evaluation of image-guidance protocols in the treatment of head and neck cancers. International Journal of Radiation Oncology* Biology* Physics. 2007;67(3):670–677.

6. Hawkins MA, Brock KK, Eccles C, Moseley D, Jaffray D, Dawson LA. Assessment of residual error in liver position using kV cone-beam computed tomography for liver cancer high-precision radiation therapy. International Journal of Radiation Oncology* Biology* Physics. 2006;66(2):610–619.

7. Meyer J, Wilbert J, Baier K, et al. Positioning accuracy of cone-beam computed tomography in combination with a HexaPOD robot treatment table. International Journal of Radiation Oncology* Biology* Physics. 2007;67(4):1220–1228.

8. Kupelian PA, Lee C, Langen KM, et al. Evaluation of image-guidance strategies in the treatment of localized prostate cancer. International Journal of Radiation Oncology* Biology* Physics. 2008;70(4):1151–1157.

9. Guckenberger M, Meyer J, Wilbert J, et al. Intra-fractional uncertainties in cone-beam CT based image-guided radiotherapy (IGRT) of pulmonary tumors. Radiotherapy and oncology. 2007;83(1):57–64.

10. Sethi R, Sandhu A, Rice R, et al. Prostate bed localization with image guided approach using On-board imaging: reporting acute toxicity, prostate bed movement, and implications for radiation therapy planning following prostatectomy. International Journal of Radiation Oncology• Biology• Physics. 2007;69(3):S341.

11. Sonke J-J, Rossi M, Wolthaus J, van Herk M, Damen E, Belderbos J. Frameless stereotactic body radiotherapy for lung cancer using four-dimensional cone beam CT guidance. International Journal of Radiation Oncology* Biology* Physics. 2009;74(2):567–574.

12. Craig T, Moiseenko V, Battista J, Van Dyk J. The impact of geometric uncertainty on hypofractionated external beam radiation therapy of prostate cancer. International Journal of Radiation Oncology* Biology* Physics. 2003;57(3):833–842.

13. Chetty IJ, Rosu M, Tyagi N, et al. A fluence convolution method to account for respiratory motion in three-dimensional dose calculations of the liver: A Monte Carlo study. Medical physics. 2003;30(7):1776–1780.

14. Song WY, Schaly B, Bauman G, Battista JJ, Van Dyk J. Evaluation of image-guided radiation therapy (IGRT) technologies and their impact on the outcomes of hypofractionated prostate cancer treatments: a radiobiologic analysis. International Journal of Radiation Oncology* Biology* Physics. 2006;64(1):289–300.

15. van Haaren PM, Bel A, Hofman P, van Vulpen M, Kotte AN, van der Heide UA. Influence of daily setup measurements and corrections on the estimated delivered dose during IMRT treatment of prostate cancer patients. Radiotherapy and oncology. 2009;90(3):291–298.

16. Velec M, Moseley JL, Eccles CL, et al. Effect of breathing motion on radiotherapy dose accumulation in the abdomen using deformable registration. International Journal of Radiation Oncology* Biology* Physics. 2011;80(1):265–272.

17. Deek MP, Kim S, Yue N, et al. Modern radiotherapy using image guidance for unresectable non-small cell lung cancer can improve outcomes in patients treated with chemoradiation therapy. Journal of thoracic disease. 2016;8(9):2602.

18. Kilburn JM, Soike MH, Lucas JT, et al. Image guided radiation therapy may result in improved local control in locally advanced lung cancer patients. Practical radiation oncology. 2016;6(3):e73–e80.

19. Du XL, Fang S, Vernon SW, et al. Racial disparities and socioeconomic status in association with survival in a large population-based cohort of elderly patients with colon cancer. Cancer. 2007;110(3):660–669.

20. https://healthcaredelivery.cancer.gov/seermedicare/program/comorbidity.html.NCIDoCCPSS-MCoCWa.

21. Charlson ME, Sax FL, MacKenzie CR, Fields SD, Braham RL, Douglas RG, Jr. Assessing illness severity: does clinical judgment work? J Chronic Dis. 1986;39(6):439–452.

22. Klabunde CN, Legler JM, Warren JL, Baldwin LM, Schrag D. A refined comorbidity measurement algorithm for claims-based studies of breast, prostate, colorectal, and lung cancer patients. Ann Epidemiol. 2007;17(8):584–590.

23. Klabunde CN, Potosky AL, Legler JM, Warren JL. Development of a comorbidity index using physician claims data. J Clin Epidemiol. 2000;53(12):1258–1267.

24. Centers for Medicare & Medicaid Services (CMS) Chronic Conditions Data Warehouse (CCW) Chronic Condition Algorithms. Definitions at https://www.ccwdata.org/web/guest/condition-categories.

25. Warren JL, Harlan LC, Fahey A, et al. Utility of the SEER-Medicare data to identify chemotherapy use. Med Care. 2002;40(8 Suppl):IV-55-61.

26. Boero IJ, Paravati AJ, Xu B, et al. Importance of radiation oncologist experience among patients with head-and-neck cancer treated with intensity-modulated radiation therapy. Journal of Clinical Oncology. 2016;34(7):684–690.

27. Shirvani SM, Jiang J, Gomez DR, Chang JY, Buchholz TA, Smith BD. Intensity modulated radiotherapy for stage III non-small cell lung cancer in the United States: predictors of use and association with toxicities. Lung cancer (Amsterdam, Netherlands). 2013;82(2):252–259.

28. Smith BD, Pan I-W, Shih Y-CT, et al. Adoption of intensity-modulated radiation therapy for breast cancer in the United States. Journal of the National Cancer Institute. 2011;103(10):798–809.

29. Chen AB. Comparative effectiveness research in radiation oncology: assessing technology. Paper presented at: Seminars in radiation oncology2014.

30. Kong F-MS. What happens when proton meets randomization: Is there a future for proton therapy? 2018.

31. Tøndel H, Lund J-Å, Lydersen S, et al. Radiotherapy for prostate cancer–Does daily image guidance with tighter margins improve patient reported outcomes compared to weekly orthogonal verified irradiation? Results from a randomized controlled trial. Radiotherapy and Oncology. 2018;126(2):229–235.

32. De Crevoisier R, Bayar MA, Pommier P, et al. Daily Versus Weekly Prostate Cancer Image Guided Radiation Therapy: Phase 3 Multicenter Randomized Trial. International Journal of Radiation Oncology* Biology* Physics. 2018;102(5):1420–1429.

