## Appendix for "Efficacy and Cost of High-Frequency IGRT in Elderly Stage III Non-Small-Cell Lung Cancer Patients"

| **Page #** | **Supplementary Section Number** | **Title** |
| --- | --- | --- |
| 2 | M1 | Definition of definitive (i.e., non-palliative) radiotherapy. |
| 2 | M2 | Motivation and validation of our claims based definition for hfIGRT. |
| 2 | M3 | Description and calculation of covariates. |
| 3 | M4 | Calculating cost of care |
| 3-4 | M5 | Using Local Coverage Determinations |
| 4-5 | M6 | Statistical analysis |
| 6 | T1 | Consort diagram describing our selection process. |
| 7 | T2 | Codes used for delineation of procedures for diagnosis and treatment. |
| 8-10 | T3 | Bivariate distribution table of patient characteristics |
| 11-13 | T4 | Codes used for delineation of radiation complications |
| 13-16 | T5 | Multivariate proportional hazards regression for lung toxicity (broad definition) |
| 17-19 | T6 | Multivariate proportional hazards regression for lung toxicity (narrow definition) |
| 20-22 | T7 | Multivariate proportional hazards regression for esophagus toxicity (all components) |
| 23-25 | T8 | Multivariate proportional hazards regression for esophagitis (based on diagnosis codes) |
| 26-28 | T9 | Multivariate proportional hazards regression for dehydration (based on diagnosis codes) |
| 29-31 | T10 | Multivariate proportional hazards regression for feeding tube placement (based on procedural codes) |
| 32-34 | T11 | Multivariate proportional hazards regression for mucositis (based on diagnosis codes) |
| 35-37 | T12 | Multivariate proportional hazards regression for cancer-specific survival |
| 38-40 | T13 | Multivariate proportional hazards regression for overall survival |
| 41-43 | T14 | Bivariate distribution table of 6-month survivor cohort used in cost analysis |
| 44 | T15 | Costs of treatment |
| 45 | F1 | Distribution of IGRT Utilization by Patient |
| 46 | F2 | Billing for Port/KV vs. CBCT Films for IGRT Over Time |
| 47 | F3 | Distribution of CBCT Utilization in Patients Who Received hfIGRT |
| 48 | F4 | Distribution of IGRT Utilization by Physician |
| 49 | F5 | IGRT Cost by LCD |
| 50 | References | References for the appendix |

**Supplementary Section M1**. Definition of definitive (i.e., non-palliative) radiotherapy.

Prescriptions for definitive radiotherapy vary by institution, physician, and patient-specific circumstances. Common – but certainly not the only – prescriptions for definitive radiation for stage III lung cancer include 6000cGy delivered in 30 once daily fractions to 6660cGy given in 37 once daily fractions. To reduce the likelihood of including patients treated with palliative intent for metastatic disease, we defined a definitive course of radiation as: (i) between 25 and 45 radiotherapy fractions; (ii) completed within 6 months of diagnosis. The number of radiotherapy fractions was calculated by counting the number of unique dates on which clusters of CPT codes for radiation delivery were billed (**Supplementary Table T1**). Because patients who recur or develop distant metastatic disease can receive additional radiation, we assumed that (iii) a break of 30 days or more between sequential radiation codes indicated an additional course of radiation. Radiation administered after a 30 day break was not included in the count of total fractions administered.

**Supplementary Section M2**. Motivation and validation of our claims based definition for hfIGRT.

To create a claims based definition of hfIGRT, we started by calculating for each patient the ratio of images-to-treatment fractions using the following formula:

$$\% imaging during treatment= \frac{\# images}{\# radiation treatment fractions}$$

An imaging-to-fraction ratio of 100% means a patient received daily imaging with each and every fraction of radiation treatment. A ratio of 0% means a patient never received any imaging with any fraction of radiation treatment.

**Supplementary** **Figure F1** shows the distribution of the ratio of imaging-to-fractions received by patients during radiation treatment. It shows two clear peaks. The first peak, centered around 20%, corresponds to patients that received imaging on one weekday per week. The second peak, centered around 100%, corresponds to patients that received imaging on each and every weekday. Less than 5% of patients were present in the nadir between the two peaks. We chose a 65% image-to-fraction cutoff for hfIGRT because, by inspection, it approximated the position of this low-point. Hence, a small change in this cutoff in the positive or negative direction would have almost no effect on the number of patients defined as receiving hfIGRT. Furthermore, a 65% cutoff clearly divides the weekly and daily IGRT groups in two.

**Supplementary Section M3**. Description and calculation of covariates.

For each patient, several additional covariates were collected or calculated. We collected patient demographic and tumor information, including age at diagnosis, gender, race, marital status, year of diagnosis, and primary tumor size, grade, and stage from SEER registry data.

Using Medicare claims in the 12 months prior to cancer diagnosis, we calculated the Klabunde adaptation of the Charlson comorbidity index to assess the prevalence of comorbid disease in our cohort.^1-4^ We separately used the Centers for Medicaid & Medicare Services (CMS) Chronic Conditions Data Warehouse (CCW) algorithms to identify additional non-Charlson scored comorbid conditions, including chronic obstructive pulmonary disease (COPD).^5^

*Radiation Treatment Characteristics*

Patients were sub-stratified according to whether or not they were treated using intensity-modulated radiation therapy (IMRT). IMRT was defined as the presence of any IMRT planning or treatment billing code during the course of radiation (**Supplementary Table T1**).

We separately reported the number of fractions delivered, whether patients received therapy in a hospital or freestanding clinic, and if they received care in a rural or urban setting based on SEER registry data.

*Cancer-related factors and non-radiation treatment*

Using Medicare MEDPAR, OUTSAF, NCH, HHA, and DME claims codes (**Supplementary Table T1**), we tabulated diagnosis and staging procedures. In a similar fashion, we recorded treatments given before, after or during radiotherapy, including oxygen, surgery, and/or chemotherapy. We classified patients as treated with intravenous chemotherapy if they received it within 6 months of cancer diagnosis.^6^

*Provider volume*

We defined provider volume as the number of patients with NSCLC treated by a physician in a year using technique developed by Boreo et. al.^7^ Existing literature has confirmed a high degree of correlation between patient volumes calculated using this technique and actual physician volume. We identified the specific provider with the older Unique Physician Identification Number (UPIN) or newer National Physician Identifier (NPI) on the weekly management code, 77247. This code has the virtue of being provider specific, unlike other technical codes that link to facilities or organizations. A crosswalk file allowed us to identify patients with both an NPI and a UPIN and prevent us from double counting them. Provider volume was expressed as patients treated per year, which was defined as the total number of patients with NSCLC divided by the time interval between the treatment dates of the first and last patient.

**Supplementary Section M4**. Calculating cost of care

Total costs included Medicare payment aggregated from inpatient facility claims in the Part A Medicare Provider Analysis and Review (MEDPAR) files, outpatient facility claims in the Part B hospital-based Outpatient Claims (OUTSAF) files, and the in- or outpatient physician claims in the Part B Carrier Claims (formerly the Physician/Supplier or NCH) files. Narrower radiotherapy and related costs summed claims based on the CPT codes between 77261 – 77999 in the Outpatient and Carrier Claims files (**Supplemental Table T1**).

We adjusted costs for inflation, normalizing them to the year 2017 using the Prospective Pricing Index for Part A claims and the Medicare Economic Index for Part B claims. We simultaneously adjusted costs for geographic variation using the geographic adjustment factor for Part A claims and the Geographic Practice Cost index for Part B claims.

The National Cancer Institute’s Health Services and Economics Branch of the Applied Research Program provided all the adjustments used to tabulate costs.

**Supplementary Section M5**. Using Local Coverage Determinations

A secondary endpoint looked at whether Carrier Local Coverage Determinations (LCDs) correlated with hfIGRT use. Medicare’s administrator, the Centers for Medicare and Medicaid (CMS), uses regional, private companies to handle certain Medicare claims and their processing. Contractors were originally called Fiscal Intermediaries for Part A claims or Carriers for Part B claims. (After passage of the Part D prescription drug amendments in 2003, CMS changed their names to A or B Medicare Administrative Contractors (MACs)). These contractors create LCDs for their coverage area that provides guidance on when it is appropriate to bill under a given CPT code. LCDs cover territories that generally, but not exactly, overlap the 16 SEER registries contained in the SEER-Medicare database.

For our study, we initially looked for – but failed to find any - LCDs governing IGRT use. Hypothesizing that IMRT and IGRT use are correlated, we next sought LCDs covering intensity-modulated radiotherapy (IMRT) billing and binned them into one of three groups as defined by Smith et al.: (1) LCDs that had restrictive IMRT guidance (that expressly permitted only inverse computer planning), (2) LCDs that had permissive IMRT guidance (that allowed both inverse and non-inverse planning), and (3) LCDs that had no IMRT guidance. But why investigate IMRT LCDs in a study of IGRT?

IMRT is a method of radiation delivery that can potentially benefit from more frequent (i.e., high-frequency) IGRT use. In contrast to non-IMRT, IMRT uses more treatment angles and computer-controlled, variable modulation of the treatment beam intensity. On the one hand, these features make IMRT radiation treatment times longer and more expensive. On the other hand, these same features generally make IMRT treatments more precise (or conformal); as a result, oncologists can in theory use much smaller planning treatment volume (PTV) margins. But using smaller PTV margins requires more accurate patient positioning, the very problem IGRT was developed to address. Hence, IGRT use should be closely correlated with IMRT use and this intuition was validated by our logistic regression. Since physicians – not hospitals – ultimately decide whether or not to use IMRT and IGRT, we focused on Carrier LCDs since they govern physician reimbursement (versus hospital reimbursement, which is managed with difference guidelines) and, by extension, shape physician incentives.

Similar to Smith et al, we identified 10 registries (Connecticut, Greater California, Hawaii, Iowa, Los Angeles, New Jersey, New Mexico post-2008, San Francisco, San Jose, Seattle, and Utah post-2007) with restrictive IMRT LCDs during our study period from 2006 – 2013; 3 registries with permissive IMRT LCDs (Detroit, Atlanta, and rural Georgia); and 3 registries (Kentucky, Louisiana, New Mexico pre-2008, and Utah pre-2007) that lacked any IMRT LCDs.

**Supplementary Section M6**. Statistical analysis

*Chi-square bivariate analysis***.** We compared the distribution of patient characteristics between the two treatment groups with the Pearson’s chi-square test.

*Logistic regression testing associations with hfIGRT utilization***.** Bivariate associations at a significance level of p = 0.20 or less were included in an initial multivariable logistic regression model to predict hfIGRT utilization. Logistic models were calculated using both state and LCD as a proxy for geographic location. The model was modified using stepwise forward and backwards elimination with threshold values of p ≤ 0.20 and p ≤ 0.05, respectively. We assessed the quality of our model by checking the area under the curve (c = 0.86 > 0.70 for the state-based model, similar for the LCD model) and the Hosmer and Lemeshow goodness of fit (p = 0.23 > 0.05 or the state-based model, similar for the LCD model).

*Kaplan-Meier analysis testing association between hfIGRT and toxicity***.** The association between hfIGRT and toxicities were assessed using the Kaplan-Meier method with censorship at the earliest of the following: death, or the end of the study period on December 31, 2013. For each endpoint, the proportional hazards assumption with respect to radiation technique was tested visually by inspection of log-log plots and analytically using Schoenfield residuals.

*Cox regression testing associations with toxicity***.** As in our logistic model, Cox regressions were performed using stepwise forward and backwards elimination with threshold values of p ≤ 0.20 and p ≤ 0.05. Clinically important covariates including age, oxygen status, performance status, stage, number of radiation treatments, and treatment strategy were included in the final model regardless of their p-values during selection. For all toxicity endpoints, the proportional hazards assumption with respect to radiation technique was satisfied, and goodness- of-fit for all final models was acceptable (p > 0.05).

*Propensity score matching***.** Based on the results of our logistic regression, we created a propensity score model to validate the findings of the multivariate Cox regressions. Patients were randomly sorted and then matched 1-to-1 without replacement to a nearest neighbor with a match caliper of 0.01. Bivariate association p-values were used to ensure that matched patients were well-balanced across covariates. Proportional hazards models, adjusted for unbalanced covariates (p < 0.20), were generated to compare the cohorts using forward and backward selection as described above. Plots of toxicity on the matched cohorts were re-generated using the Kaplan-Meier method.

*Survival analysis***.** Overall and cause-specific survival was tested in the same way as toxicities with multivariate Cox regressions and matched cohort analysis.

To protect patient anonymity and consistent with policies governing the use of SEER-Medicare data, none of the tables report the number of patients in any sub-group with a sample size less than 11. Analyses were performed with SAS 9.4 (SAS, Cary, NC).

*Estimating the cost differential between the two treatment groups*. A non-parametric bootstrap model used 1,000 samples to estimate the 95% confidence interval (CI) around the mean cost difference between the hfIGRT and non-hfIGRT groups. We estimated these differences for both the original, unmatched cohort and the 1-to-1 matched sub-cohort.

*Wilcoxon bivariate analysis of covariates against cost*. The impact of individual (categorical) covariates on the (continuous) cost of treatment was assessed using the Wilcoxon two-sample test.

To protect patient anonymity and consistent with policies governing the use of SEER-Medicare data, none of the tables report the number of patients in any sub-group with a sample size less than 11. Analyses were performed with SAS 9.4 (SAS, Cary, NC).

| **Supplementary Table T1**. Consort diagram describing our selection process. | | |
| --- | --- | --- |
| **Selection Criteria** | **Number of Remaining Obs.** | |
|  | **hfIGRT** | **No hfIGRT** |
| All Lung Cancer Patients in the SEER-Medicare Database | 600,828 | |
| Only Select SEER-Records for First Cancer Diagnosis | 548,939 | |
| 1st Cancer is of the Lung | 471,916 | |
| Reporting Source should not be autopsy or death certificate. | 459,716 | |
| Age of diagnosis should be greater than 65. | 381,444 | |
| Original or current reason for entitlement should be age. | 379,047 | |
| Delete if date of death between SEER and Medicare if off by > 3 months. | 378,158 | |
| Take only cases diagnosed from 2006 to 2011 | 141,552 | |
| Exclude members of an HMO 12 months before to 12 months after diagnosis. | 102,101 | |
| Have both Part A & B Coverage | 89,680 | |
| Non-small cell lung cancer | 64,949 | |
| Stage III | 15,874 | |
| Definitive Treatment: 25 to 45 Radiation Fractions | 5,157 | |
| Cohort | 962 | 3468 |

**Table T1**: Consort diagram showing selection criteria and number of patients left in cohort after each selection criteria is applied.

| **Supplementary Table T2**. Codes used for delineation of procedures for diagnosis and treatment | | | | |
| --- | --- | --- | --- | --- |
| **Treatment** | **ICD-9 Codes** | **CPT/HCPCS Codes** | **Revenue Center/**  **Diagnosis Related Codes** | **DME File MTUSIND Code** |
| Chemotherapy | 99.25,  V58.1, V66.2, V67.2  E9331, E9307 | Q0083, Q0084, Q0085,  96400-96599,  J9000-J9999  G0355-G0362 | 0331, 0332, 0335  410 |  |
| Cone-beam CT (CBCT) |  | 77014, 77421 |  |  |
| Portal/KV Films |  | 77417 |  |  |
| Image-guided Radiation Therapy (IGRT = CBCT + Port/KV) |  | 77014, 77421, 77417 |  |  |
| Intensity-modulate Radiation Therapy (IMRT) |  | 77301, 77338, 77418, G0174, G0178 |  |  |
| PET staging scan |  | 78810-78816  G0030-G0047  G0210-G0235  G0125-G0126  G0163-G0165  G0252-G0254  G0296, G0330, G0331, G0336 |  |  |
| Radiation | V58.0, V66.1, V67.1  92.21,92.22,92.23,92.24,92.25,92.26,92.27,92.28,92.29 | 77401-77499,  77520-77525,  77750-77799 | 0330, 0333 |  |
| Supplemental O2 |  |  |  | 4 |
| Surgery | 321, 323, 324, 325, 326, 3220, 3229, 3230, 3239, 3241, 3249, 3250, 3259 | 32440, 32442, 32445, 32480, 32482, 32484, 32486, 32488, 32500, 32503, 32504, 32520, 32522, 32525, 32657, 32663 |  |  |
| Abbreviations: ICD-9, International Classification of Diseases, 9th Revision, Clinical Modification (ICD-9-CM); CPT/HCPCS, Current Procedural Terminology/ Healthcare Common Procedure Coding System | | | | |

| **Supplementary Table T3**. Bivariate distribution table of patient characteristics | | | | | | | | | | |
| --- | --- | --- | --- | --- | --- | --- | --- | --- | --- | --- |
| Characteristic | ***Unmatched*** | | | | | ***Matched*** | | | | |
|  | IGRT (N) | IGRT (%) | No IGRT (N) | No IGRT (%) | P-value | IGRT (N) | IGRT (%) | No IGRT (N) | No IGRT (%) | P-value |
| Age |  |  |  |  | 0.41 |  |  |  |  | 0.80 |
| 65 - 74 | 508 | 52.8 | 1911 | 55.1 |  | 364 | 51.1 | 376 | 52.7 |  |
| 75 - 84 | 397 | 41.3 | 1349 | 38.9 |  | 306 | 42.9 | 294 | 41.2 |  |
| 85+ | 57 | 5.9 | 208 | 6 |  | 43 | 6 | 43 | 6 |  |
| Sex |  |  |  |  | 0.89 |  |  |  |  | 0.29 |
| Male | 528 | 54.9 | 1895 | 54.6 |  | 394 | 55.3 | 374 | 52.5 |  |
| Female | 434 | 45.1 | 1573 | 45.4 |  | 319 | 44.7 | 339 | 47.5 |  |
| Race |  |  |  |  | 0.07 |  |  |  |  | 0.05 |
| White | ≥838 | ≥87.2 | 2979 | 85.9 |  | ≥632 | ≥88.3 | ≥619 | ≥86.8 |  |
| Black | 77 | 8 | 308 | 8.9 |  | 51 | 7.2 | 54 | 7.6 |  |
| Hispanic | ≤11 | ≤1.1 | 33 | 1 |  | ≤11 | ≤1.5 | ≤11 | ≤1.5 |  |
| Other | 35 | 3.6 | 148 | 4.3 |  | 19 | 2.7 | 29 | 4.1 |  |
| State |  |  |  |  | <.01 |  |  |  |  | 0.11 |
| California | ≥178 | ≥18.6 | 777 | 22.4 |  | ≥100 | ≥15.4 | ≥146 | ≥21.3 |  |
| Connecticut | 54 | 5.6 | 254 | 7.3 |  | 54 | 7.6 | 42 | 5.9 |  |
| Georgia | 145 | 15.1 | 507 | 14.6 |  | 93 | 13.0 | 108 | 15.1 |  |
| Hawaii | ≤11 | ≤1.1 | 45 | 1.3 |  | ≤11 | ≤1.5 | ≤11 | ≤1.5 |  |
| Iowa | 76 | 7.9 | 228 | 6.6 |  | 75 | 10.5 | 50 | 7 |  |
| Kentucky | 101 | 10.5 | 368 | 10.6 |  | 76 | 10.7 | 68 | 9.5 |  |
| Louisiana | 70 | 7.3 | 285 | 8.2 |  | 63 | 8.8 | 53 | 7.4 |  |
| Michigan | 83 | 8.6 | 264 | 7.6 |  | 80 | 11.2 | 71 | 10 |  |
| New Jersey | 142 | 14.8 | 459 | 13.2 |  | 81 | 11.4 | 81 | 11.4 |  |
| New Mexico | ≤11 | ≤1.1 | 63 | 1.8 |  | ≤11 | ≤1.5 | ≤11 | ≤1.5 |  |
| Utah | ≤11 | ≤1.1 | 30 | 0.9 |  | ≤11 | ≤1.5 | ≤11 | ≤1.5 |  |
| Washington | 80 | 8.3 | 188 | 5.4 |  | 58 | 8.1 | 61 | 8.6 |  |
| Local Coverage Determination |  |  |  |  | 0.62 |  |  |  |  | 0.48 |
| Favorable | 220 | 22.9 | 750 | 21.6 |  | 170 | 23.8 | 177 | 24.8 |  |
| Intermediate | 562 | 58.4 | 2033 | 58.6 |  | 395 | 55.4 | 406 | 56.9 |  |
| Unfavorable | 180 | 18.7 | 685 | 19.8 |  | 148 | 20.8 | 130 | 18.2 |  |
| Year of Diagnosis |  |  |  |  | <.01 |  |  |  |  | 0.85 |
| 2006 | 14 | 1.5 | 785 | 22.6 |  | 14 | 2 | 12 | 1.7 |  |
| 2007 | 71 | 7.4 | 741 | 21.4 |  | 71 | 10 | 73 | 10.2 |  |
| 2008 | 103 | 10.7 | 663 | 19.1 |  | 101 | 14.2 | 105 | 14.7 |  |
| 2009 | 202 | 21 | 534 | 15.4 |  | 157 | 22 | 142 | 19.9 |  |
| 2010 | 241 | 25.1 | 402 | 11.6 |  | 175 | 24.5 | 193 | 27.1 |  |
| 2011 | 331 | 34.4 | 343 | 9.9 |  | 195 | 27.3 | 188 | 26.4 |  |
| Charlson Score (No COPD) |  |  |  |  | 0.16 |  |  |  |  | 0.44 |
| 0 | ≥503 | ≥52.3 | 1952 | 56.3 |  | ≥372 | ≥52.2 | ≥397 | ≥55.7 |  |
| 1-2 | 359 | 37.3 | 1199 | 34.6 |  | 267 | 37.4 | 245 | 34.4 |  |
| > 2 | 95 | 9.9 | 317 | 9.1 |  | 67 | 9.4 | 66 | 9.3 |  |
| COPD | 457 | 47.5 | 1590 | 45.8 | 0.36 | 329 | 46.1 | 330 | 46.3 | 0.96 |
| Supplemental O2 | 234 | 24.3 | 864 | 24.9 | 0.71 | 170 | 23.8 | 176 | 24.7 | 0.71 |
| Homebound | ≤11 | ≤1.1 | 67 | 1.9 | <.01 | ≤11 | ≤1.5 | ≤11 | ≤1.5 | 0.53 |
| Histology |  |  |  |  | <.01 |  |  |  |  | 0.18 |
| Adenocarcinoma | 377 | 39.2 | 1086 | 31.3 |  | 276 | 38.7 | 237 | 33.2 |  |
| SCC | 397 | 41.3 | 1463 | 42.2 |  | 295 | 41.4 | 314 | 44 |  |
| Large Cell | 23 | 2.4 | 114 | 3.3 |  | 19 | 2.7 | 21 | 2.9 |  |
| Other | 165 | 17.2 | 805 | 23.2 |  | 123 | 17.3 | 141 | 19.8 |  |
| Stage |  |  |  |  | 0.37 |  |  |  |  | 0.53 |
| Stage IIIA | 489 | 50.8 | 1706 | 49.2 |  | 366 | 51.3 | 354 | 49.6 |  |
| Stage IIIB | 473 | 49.2 | 1762 | 50.8 |  | 347 | 48.7 | 359 | 50.4 |  |
| T-Stage |  |  |  |  | 0.74 |  |  |  |  | 0.97 |
| TX | 47 | 4.9 | 148 | 4.3 |  | 33 | 4.6 | 30 | 4.2 |  |
| T0 | ≤11 | ≤1.1 | 18 | 0.5 |  | ≤11 | ≤1.5 | ≤11 | ≤1.5 |  |
| T1 | 132 | 13.7 | 471 | 13.6 |  | 101 | 14.2 | 108 | 15.1 |  |
| T2 | 326 | 33.9 | 1135 | 32.7 |  | 235 | 33.0 | 227 | 31.8 |  |
| T3 | 96 | 10 | 327 | 9.4 |  | 72 | 10.1 | 70 | 9.8 |  |
| T4 | ≥350 | ≥36.4 | 1369 | 39.5 |  | ≥261 | ≥36.6 | ≥267 | ≥37.5 |  |
| Tumor Size |  |  |  |  | 0.54 |  |  |  |  | 0.95 |
| < 2.0 | 62 | 6.4 | 246 | 7.1 |  | 42 | 5.9 | 45 | 6.3 |  |
| 2.0-5.0 | 435 | 45.2 | 1589 | 45.8 |  | 329 | 46.1 | 319 | 44.7 |  |
| > 5.0 | 328 | 34.1 | 1105 | 31.9 |  | 245 | 34.4 | 249 | 34.9 |  |
| Unknown | 137 | 14.2 | 528 | 15.2 |  | 97 | 13.6 | 100 | 14 |  |
| Tumor Laterality |  |  |  |  | 0.83 |  |  |  |  | 0.51 |
| Right | ≥556 | ≥57.8 | ≥2007 | ≥57.8 |  | ≥403 | ≥57.2 | ≥387 | ≥54.8 |  |
| Left | 383 | 39.8 | 1416 | 40.8 |  | 288 | 40.4 | 304 | 42.6 |  |
| Unpaired | ≤11 | ≤1.1 | ≤11 | 0.3 |  | ≤11 | ≤1.5 | ≤11 | ≤1.5 |  |
| Unknown | 12 | 1.2 | 34 | 1 |  | ≤11 | ≤1.5 | ≤11 | ≤1.5 |  |
| Tumor Location |  |  |  |  | 0.15 |  |  |  |  | 0.08 |
| Main bronchus | 52 | 5.4 | 223 | 6.4 |  | 35 | 4.9 | 56 | 7.9 |  |
| Upper lobe | ≥573 | ≥59.6 | 2023 | 58.3 |  | ≥422 | ≥59.2 | ≥420 | ≥58.9 |  |
| Middle lobe | 27 | 2.8 | 138 | 4 |  | 20 | 2.8 | 19 | 2.7 |  |
| Lower lobe | 243 | 25.3 | 872 | 25.1 |  | 183 | 25.7 | 173 | 24.3 |  |
| Lung NOS | 56 | 5.8 | 182 | 5.2 |  | 42 | 5.9 | 34 | 4.8 |  |
| Other | ≤11 | ≤1.1 | 30 | 0.9 |  | ≤11 | ≤1.5 | ≤11 | ≤1.5 |  |
| PET Staging | 902 | 93.8 | 3194 | 92.1 | 0.08 | 664 | 93.1 | 659 | 92.4 | 0.61 |
| # of nodes positive |  |  |  |  | 0.63 |  |  |  |  | 0.81 |
| 0 | 40 | 4.2 | 167 | 4.8 |  | 26 | 3.6 | 22 | 3.1 |  |
| 1-3 | 126 | 13.1 | 432 | 12.5 |  | 85 | 11.9 | 79 | 11.1 |  |
| 4+ | 25 | 2.6 | 109 | 3.1 |  | 18 | 2.5 | 22 | 3.1 |  |
| Unknown | 771 | 80.1 | 2760 | 79.6 |  | 584 | 81.9 | 590 | 82.7 |  |
| Treatment Type |  |  |  |  | 0.23 |  |  |  |  | 0.45 |
| Trimodality | 71 | 7.4 | 289 | 8.3 |  | 54 | 7.6 | 52 | 7.3 |  |
| Chemotherapy & radiation | 735 | 76.4 | 2534 | 73.1 |  | 530 | 74.3 | ≥550 | ≥77.2 |  |
| Surgery & radiation | 16 | 1.7 | 65 | 1.9 |  | 14 | 2 | ≤11 | ≤1.5 |  |
| Radiation alone | 140 | 14.6 | 580 | 16.7 |  | 115 | 16.1 | 101 | 14 |  |
| # of RT Fractions |  |  |  |  | <.01 |  |  |  |  | 0.92 |
| 25 - 29 | 196 | 20.4 | 678 | 19.6 |  | 141 | 19.8 | 137 | 19.2 |  |
| 30 - 34 | 361 | 37.5 | 1558 | 44.9 |  | 280 | 39.3 | 277 | 38.8 |  |
| 35 - 40 | 405 | 42.1 | 1232 | 35.5 |  | 292 | 41 | 299 | 41.9 |  |
| IMRT | 531 | 55.2 | 487 | 14 | <0.01 | 298 | 41.8 | 308 | 43.2 | 0.59 |
| Type of Treatment Center |  |  |  |  | <.01 |  |  |  |  | 0.25 |
| Free Standing | 401 | 41.7 | 1092 | 31.5 |  | 261 | 36.6 | 290 | 40.7 |  |
| Hospital Based | ≥550 | ≥57.2 | 2350 | 67.8 |  | ≥441 | ≥61.9 | ≥412 | ≥57.8 |  |
| Both | ≤11 | ≤1.1 | 26 | 0.7 |  | ≤11 | ≤1.5 | ≤11 | ≤1.5 |  |
| Rural vs. Urban |  |  |  |  | 0.06 |  |  |  |  | 0.50 |
| Rural | 161 | 16.7 | 674 | 19.4 |  | 141 | 19.8 | 131 | 18.4 |  |
| Urban | 801 | 83.3 | 2794 | 80.6 |  | 572 | 80.2 | 582 | 81.6 |  |
| Radiation Oncologist Density |  |  |  |  | <.01 |  |  |  |  | 0.31 |
| 1st quartile | 285 | 29.6 | 1131 | 32.6 |  | ≥201 | ≥28.2 | ≥197 | ≥27.6 |  |
| 2nd quartile | ≥287 | ≥29.9 | 893 | 25.7 |  | 194 | 27.2 | 226 | 31.8 |  |
| 3rd quartile | 221 | 23 | 788 | 22.7 |  | 188 | 26.4 | 176 | 24.7 |  |
| 4th quartile | 158 | 16.4 | 611 | 17.6 |  | 119 | 16.7 | 103 | 14.4 |  |
| Unknown | ≤11 | ≤1.1 | 45 | 1.3 |  | ≤11 | ≤1.5 | ≤11 | ≤1.5 |  |
| General Surgeon Density |  |  |  |  | <.01 |  |  |  |  | 0.21 |
| 1st quartile | 252 | 26.2 | 1095 | 31.6 |  | 194 | 27.3 | 198 | 27.8 |  |
| 2nd quartile | 234 | 24.3 | 804 | 23.2 |  | 155 | 21.7 | 178 | 25 |  |
| 3rd quartile | ≥248 | ≥26.2 | 821 | 23.7 |  | ≥192 | ≥26.9 | ≥199 | ≥28 |  |
| 4th quartile | 217 | 22.6 | 703 | 20.3 |  | 161 | 22.6 | 127 | 17.8 |  |
| Unknown | ≤11 | ≤1.1 | 45 | 1.3 |  | ≤11 | ≤1.5 | ≤11 | ≤1.5 |  |
| Physician Experience |  |  |  |  | <.01 |  |  |  |  | 0.13 |
| 1st quartile | 173 | 18 | ≥928 | ≥26.7 |  | 145 | 20.3 | 150 | 21 |  |
| 2nd quartile | 244 | 25.4 | 861 | 24.8 |  | 184 | 25.8 | 216 | 30.3 |  |
| 3rd quartile | 335 | 34.8 | 772 | 22.3 |  | 217 | 30.4 | 183 | 25.7 |  |
| 4th quartile | 210 | 21.8 | 896 | 25.8 |  | 167 | 23.4 | 164 | 23 |  |
| Other | 0 | 0 | ≥11 | 0.3 |  | 0 | 0 | 0 | 0 |  |
| * To protect patient anonymity, all cells with values between 1 and 11 were masked with the indicator “≤11.” Then the largest cell in the same column for the same characteristic was adjusted in the opposite direction so that the total number of observations in that column remained the same. | | | | | | | | | | |

| **Supplementary Table T4**. Codes used for delineation of radiation complications | | | | |
| --- | --- | --- | --- | --- |
| **Pulmonary Complications:** | | | | |
| **Narrow Acute Lung Toxicity** | | | |  |
|  | ICD-9 | | 508.0 | Acute pulmonary manifestations due to radiation |
| **Broad Acute Long Toxicity** | | | |  |
|  | ICD-9 | | 508.0 | Acute pulmonary manifestations due to radiation |
|  |  | | 485 | Bronchopneumonia, organism unspecified |
|  |  | | 486 | Pneumonia, organism unspecified |
|  |  | | 482.9 | Bacterial pneumonia, unspecified |
|  |  | | 514 | Pulmonary congestion and hypostasis (NOT attributable to heart failure) |
| **Esophageal Complications:** | | | | |
| **Esophagitis and Dysphagia** | | | |  |
|  | ICD-9 | 530.1 | | Esophagitis |
|  |  | 530.11 | | Esophagitis, unspecified |
|  |  | 530.12 | | Esophagitis, acute |
|  |  | 530.19 | | Esophagitis, other types |
|  |  | 787.2 | | Dysphagia |
|  |  | 787.20 | | Dysphagia unspecified |
|  |  | 787.21 | | Dysphagia oral phase |
|  |  | 787.22 | | Dysphagia oropharyngeal phase |
|  |  | 787.23 | | Dysphagia pharyngeal phase |
|  |  | 787.24 | | Dysphagia pharyngoesophageal phase |
|  |  | 787.29 | | Other dysphagia |
|  |  | 784.1 | | Throat pain |
| **Dehydration** | | | |  |
|  | ICD-9 | 276.5 | | Volume depletion |
|  |  | 276.50 | | Volume depletion, unspecified |
|  |  | 276.51 | | Dehydration |
|  |  | 276.52 | | Hypovolemia |
| **Mucositis** | |  | |  |
|  | ICD-9 | 528.0 | | Stomatitis and mucositis |
|  |  | 528.00 | | Mucositis, unspecified |
|  |  | 528.01 | | Mucositis (ulcerative) due to antineoplastic therapy |
|  |  | 528.02 | | Mucositis (ulcerative) due to other drug |
|  |  | 528.09 | | Mucositis (ulcerative) due to other cause |
|  |  | 538 | | Gastrointestinal mucositis |
| **Feeding Tube Placement** | | | |  |
|  | ICD-9 | 43.1 | | Gastrostomy |
|  |  | 43.11 | | Percutaneous enterogastrostomy |
|  |  | 43.19 | | Other gastrostomy |
|  |  | 46.3 | | Other enterostomy |
|  |  | 46.31 | | Delayed opening of other enterostomy |
|  |  | 46.32 | | Percutanoues enterojejunostomy |
|  |  | 46.39 | | Feeding enterostomy or Duodenostomy |
|  | CPT | 43246 | | Upper GI endoscopy with directed placement of percutaneous gastrostomy tube |
|  |  | 43750 | | Percutaneous placement of gastrostomy tube |
|  |  | 43760 | | Change of gastrostomy tube |
|  |  | 43761 | | Repositioning of gastric feedint tube through the duodenum for enteric nutrition |
|  |  | 74350 | | Percutaneous placement of gastrostomy tube, radiological supervision/interpretation |
|  |  | 74355 | | Percutaneous placement of enteroclysis tube, radiological supervision/interpretation |
|  |  | B4086 | | Gastrostomy/jejunostomy tube, any material |
|  |  | 44373 | | Small intestinal endoscopy with conversion of PEG tube to PEJ tube |
|  |  | 44372 | | Small intestinal endoscopy with placement of PEJ tube |
|  |  | 44015 | | Tube or needle catheter jejunostomy for enteral alimentation |
|  |  | 44201 | | Surgical laparoscopy with jejunostomy for feeding or nutrition |
| Abbreviations: ICD-9, International Classification of Diseases, 9th Revision, Clinical Modification (ICD-9-CM); CPT, Current Procedural Terminology/ Healthcare Common Procedure Coding System | | | | |

| **Supplementary Table T4**. Multivariable logistic regression predicting hfIGRT use, using LCD for location | | | |
| --- | --- | --- | --- |
| Predictor | Odds Ratio | 95% CI | P-value |
| Year of Diagnosis |  |  |  |
| 2006 | 1 (Ref) |  |  |
| 2007 | 6.02 | 3.32 - 10.91 | <.01 |
| 2008 | 8.35 | 4.66 - 14.95 | 0.03 |
| 2009 | 18.37 | 10.40 - 32.45 | <.01 |
| 2010 | 31.72 | 17.98 - 55.99 | <.01 |
| 2011 | 50.18 | 28.47 - 88.44 | <.01 |
| LCD |  |  |  |
| Favorable | 1 (Ref) |  |  |
| Intermediate | 1.05 | 0.83 - 1.34 | <.01 |
| Unfavorable | 0.66 | 0.50 - 0.88 | <.01 |
| General Surgeon Density |  |  |  |
| 1st quartile | 1 (Ref) |  |  |
| 2nd quartile | 1.08 | 0.81 - 1.46 | 0.02 |
| 3rd quartile | 1.53 | 1.12 - 2.09 | 0.09 |
| 4th quartile | 1.82 | 1.29 - 2.58 | <.01 |
| Unknown | - | - | - |
| Radiation Oncologist Density |  |  |  |
| 1st quartile | 1 (Ref) |  |  |
| 2nd quartile | 0.84 | 0.63 - 1.14 | <.01 |
| 3rd quartile | 0.66 | 0.48 - 0.91 | 0.12 |
| 4th quartile | 0.67 | 0.46 - 0.96 | 0.14 |
| Physician Experience |  |  |  |
| 1st quartile | 1 (Ref) |  |  |
| 2nd quartile | 2.12 | 1.64 - 2.74 | 0.33 |
| 3rd quartile | 3.45 | 2.68 - 4.44 | <.01 |
| 4th quartile | 2.06 | 1.57 - 2.70 | 0.60 |
| Type of Treatment Center |  |  |  |
| Free Standing | 1 (Ref) |  |  |
| Hospital Based | 0.63 | 0.53 - 0.76 | 0.18 |
| Both | 0.13 | 0.03 - 0.66 | 0.03 |
| # of Radiation Fractions |  |  |  |
| 25 - 29 | 1 (Ref) |  |  |
| 30 - 34 | 0.68 | 0.54 - 0.86 | <.01 |
| 35 - 40 | 0.93 | 0.74 - 1.18 | 0.19 |
| IMRT | 6.47 | 5.38 - 7.80 | <.01 |

| **Supplementary Table T5**. Multivariate proportional hazards regression for lung toxicity (broad definition).^X^ | | |
| --- | --- | --- |
| Parameter | Univariate  HR (95% CI, P-value) | Multivariate  HR (95% CI, P-Value) |
| Daily IGRT |  |  |
| No | Reference | Reference |
| Yes | 0.96 (0.86 - 1.08, 0.53) | 0.97 (0.86 - 1.09, 0.60) |
| Age |  |  |
| 65 - 74 | Reference | Reference |
| 75 - 84 | 1.09 (0.98 - 1.20, 0.10) | 1.05 (0.95 - 1.17, 0.31) |
| 85+ | 1.01 (0.82 - 1.25, 0.90) | 0.99 (0.80 - 1.23, 0.94) |
| Race |  |  |
| White | Reference | Reference |
| Black | 0.98 (0.82 - 1.15, 0.78) | * |
| Hispanic | 0.98 (0.57 - 1.69, 0.94) | * |
| Other | 0.98 (0.77 - 1.24, 0.85) | * |
| COPD |  |  |
| No | Reference | Reference |
| Yes | 1.42 (1.29 - 1.56, <.01) | 1.26 (1.13 - 1.39, <.01) |
| Charlson Score (no COPD) |  |  |
| 0 | Reference | Reference |
| 1-2 | 1.22 (1.10 - 1.35, <.01) | 1.14 (1.02 - 1.26, 0.02) |
| > 2 | 1.56 (1.34 - 1.82, <.01) | 1.41 (1.21 - 1.66, <.01) |
| Supplemental O2 |  |  |
| No | Reference | Reference |
| Yes | 1.48 (1.33 - 1.64, <.01) | 1.21 (1.08 - 1.35, <.01) |
| Homebound |  |  |
| No | Reference | Reference |
| Yes | 1.82 (1.33 - 2.49, <.01) | 1.39 (1.01 - 1.92, 0.04) |
| Stage |  |  |
| Stage IIIA | Reference | Reference |
| Stage IIIB | 1.14 (1.03 - 1.25, <.01) | 1.08 (0.98 - 1.19, 0.11) |
| T-Stage |  |  |
| TX | Reference | Reference |
| T0 | 0.65 (0.30 - 1.41, 0.27) | * |
| T1 | 0.72 (0.56 - 0.93, 0.01) | * |
| T2 | 0.88 (0.70 - 1.11, 0.28) | * |
| T3 | 1.03 (0.80 - 1.34, 0.80) | * |
| T4 | 0.98 (0.78 - 1.23, 0.85) | * |
| Tumor Size |  |  |
| < 2.0 | Reference | Reference |
| 2.0-5.0 | 1.15 (0.94 - 1.41, 0.17) | 1.11 (0.90 - 1.37, 0.32) |
| > 5.0 | 1.28 (1.04 - 1.57, 0.02) | 1.21 (0.98 - 1.50, 0.07) |
| Unknown | 1.52 (1.22 - 1.90, <.01) | 1.38 (1.10 - 1.74, <.01) |
| Histology |  |  |
| Adenocarcinoma | Reference | Reference |
| SCC | 1.26 (1.13 - 1.41, <.01) | 1.14 (1.02 - 1.28, 0.02) |
| Large Cell | 1.41 (1.09 - 1.82, <.01) | 1.28 (0.99 - 1.66, 0.06) |
| Other | 0.99 (0.86 - 1.13, 0.83) | 0.90 (0.79 - 1.03, 0.14) |
| Laterality |  |  |
| Right | Reference | Reference |
| Left | 0.96 (0.87 - 1.05, 0.37) | * |
| Unpaired | 0.77 (0.19 - 3.09, 0.72) | * |
| Unknown | 0.79 (0.47 - 1.32, 0.37) | * |
| Tumor Location |  |  |
| Main bronchus | Reference | Reference |
| Upper lobe | 0.73 (0.60 - 0.89, <.01) | 0.87 (0.72 - 1.07, 0.18) |
| Middle lobe | 0.87 (0.64 - 1.17, 0.35) | 1.02 (0.75 - 1.39, 0.90) |
| Lower lobe | 1.09 (0.89 - 1.33, 0.42) | 1.26 (1.02 - 1.55, 0.03) |
| Lung NOS | 0.84 (0.64 - 1.11, 0.23) | 0.85 (0.64 - 1.12, 0.25) |
| Other | 1.47 (0.90 - 2.42, 0.13) | 1.86 (1.12 - 3.07, 0.02) |
| PET |  |  |
| No | Reference | Reference |
| Yes | 1.00 (0.83 - 1.20, 1.00) | * |
| # of Positive Nodes |  |  |
| 0 | Reference | Reference |
| 1-3 | 0.85 (0.65 - 1.11, 0.23) | * |
| 4+ | 0.77 (0.52 - 1.12, 0.17) | * |
| Unknown | 1.16 (0.92 - 1.46, 0.20) | * |
| Treatment Type |  |  |
| Trimodality | Reference | Reference |
| Chemotherapy & radiation | 1.40 (1.15 - 1.70, <.01) | 1.54 (1.25 - 1.90, <.01) |
| Surgery & radiation | 1.06 (0.69 - 1.63, 0.78) | 1.03 (0.67 - 1.58, 0.90) |
| Radiation alone | 1.54 (1.24 - 1.92, <.01) | 1.52 (1.20 - 1.93, <.01) |
| # of RT Fractions |  |  |
| 25 - 29 | Reference | Reference |
| 30 - 34 | 0.74 (0.65 - 0.84, <.01) | 0.66 (0.58 - 0.75, <.01) |
| 35 - 40 | 0.72 (0.64 - 0.82, <.01) | 0.64 (0.56 - 0.73, <.01) |
| Type of Treatment Center |  |  |
| Free Standing | Reference | Reference |
| Hospital Based | 1.00 (0.90 - 1.10, 0.96) | * |
| Both | 1.37 (0.80 - 2.32, 0.25) | * |
| Rural vs. Urban |  |  |
| Rural | Reference | Reference |
| Urban | 0.93 (0.83 - 1.05, 0.27) | * |
| Radiation Oncologist Density |  |  |
| 1st quartile | Reference | Reference |
| 2nd quartile | 0.98 (0.87 - 1.11, 0.73) | * |
| 3rd quartile | 0.99 (0.88 - 1.13, 0.93) | * |
| 4th quartile | 0.90 (0.78 - 1.04, 0.15) | * |
| Unknown | 0.99 (0.62 - 1.58, 0.96) | * |
| General Surgeon Density |  |  |
| 1st quartile | Reference | Reference |
| 2nd quartile | 0.98 (0.86 - 1.11, 0.72) | * |
| 3rd quartile | 1.01 (0.89 - 1.14, 0.91) | * |
| 4th quartile | 1.00 (0.87 - 1.14, 0.96) | * |
| Unknown | 1.01 (0.63 - 1.61, 0.97) | * |
| Physician Experience |  |  |
| 1st quartile | Reference | Reference |
| 2nd quartile | 0.98 (0.86 - 1.12, 0.74) | * |
| 3rd quartile | 0.91 (0.79 - 1.04, 0.15) | * |
| 4th quartile | 0.96 (0.84 - 1.09, 0.50) | * |
| State |  |  |
| California | Reference | Reference |
| Connecticut | 0.89 (0.72 - 1.11, 0.31) | * |
| Georgia | 0.92 (0.78 - 1.09, 0.34) | * |
| Hawaii | 1.06 (0.66 - 1.70, 0.81) | * |
| Iowa | 1.08 (0.88 - 1.33, 0.46) | * |
| Kentucky | 1.08 (0.91 - 1.29, 0.38) | * |
| Louisiana | 1.31 (1.09 - 1.58, <.01) | * |
| Michigan | 1.09 (0.90 - 1.33, 0.37) | * |
| New Jersey | 1.10 (0.93 - 1.29, 0.26) | * |
| New Mexico | 1.03 (0.70 - 1.52, 0.89) | * |
| Utah | 1.70 (1.07 - 2.69, 0.02) | * |
| Washington | 1.01 (0.82 - 1.26, 0.91) | * |
| Year of Diagnosis |  |  |
| 2006 | Reference | Reference |
| 2007 | 0.97 (0.84 - 1.13, 0.72) | * |
| 2008 | 0.90 (0.77 - 1.06, 0.20) | * |
| 2009 | 0.91 (0.78 - 1.06, 0.24) | * |
| 2010 | 0.83 (0.70 - 0.98, 0.03) | * |
| 2011 | 0.86 (0.73 - 1.02, 0.08) | * |
| IMRT |  |  |
| No | Reference | Reference |
| Yes | 1.07 (0.96 - 1.19, 0.25) | * |
| ^X^ Multivariate Cox regressions were performed using stepwise forward and backwards elimination with threshold values of p ≤ 0.20 and p ≤ 0.05, respectively.  * Covariate auto-excluded from model during forward or backward selection.  Abbrev: HR, hazard ratio. CI, confidence interval. | | |

| **Supplementary Table T6**. Multivariate proportional hazards regression for lung toxicity (narrow definition).^X^ | | |
| --- | --- | --- |
| Parameter | Univariate  HR (95% CI, P-value) | Multivariate  HR (95% CI, P-Value) |
| Daily IGRT |  |  |
| No | Reference | Reference |
| Yes | 0.82 (0.63 - 1.06, 0.13) | 0.81 (0.62 - 1.05, 0.11) |
| Age |  |  |
| 65 - 74 | Reference | Reference |
| 75 - 84 | 1.11 (0.90 - 1.36, 0.35) | 1.04 (0.84 - 1.29, 0.71) |
| 85+ | 1.09 (0.70 - 1.70, 0.70) | 1.11 (0.69 - 1.77, 0.67) |
| Race |  |  |
| White | Reference | Reference |
| Black | 0.83 (0.56 - 1.22, 0.34) | * |
| Hispanic | 0.98 (0.31 - 3.05, 0.97) | * |
| Other | 1.01 (0.61 - 1.68, 0.95) | * |
| COPD |  |  |
| No | Reference | Reference |
| Yes | 1.05 (0.86 - 1.29, 0.63) | * |
| Charlson Score (no COPD) |  |  |
| 0 | Reference | Reference |
| 1-2 | 1.02 (0.82 - 1.26, 0.87) | * |
| > 2 | 1.00 (0.70 - 1.43, 1.00) | * |
| Supplemental O2 |  |  |
| No | Reference | Reference |
| Yes | 0.98 (0.78 - 1.24, 0.88) | 1.01 (0.79 - 1.28, 0.96) |
| Homebound |  |  |
| No | Reference | Reference |
| Yes | 1.28 (0.61 - 2.71, 0.52) | 1.15 (0.54 - 2.47, 0.71) |
| Stage |  |  |
| Stage IIIA | Reference | Reference |
| Stage IIIB | 0.94 (0.77 - 1.16, 0.58) | 0.93 (0.76 - 1.14, 0.48) |
| T-Stage |  |  |
| TX | Reference | Reference |
| T0 | 0.72 (0.17 - 3.05, 0.65) | * |
| T1 | 0.73 (0.44 - 1.20, 0.21) | * |
| T2 | 0.78 (0.49 - 1.22, 0.27) | * |
| T3 | 0.94 (0.56 - 1.56, 0.81) | * |
| T4 | 0.67 (0.42 - 1.05, 0.08) | * |
| Tumor Size |  |  |
| < 2.0 | Reference | Reference |
| 2.0-5.0 | 1.11 (0.73 - 1.68, 0.64) | * |
| > 5.0 | 1.02 (0.66 - 1.57, 0.93) | * |
| Unknown | 1.30 (0.82 - 2.06, 0.26) | * |
| Histology |  |  |
| Adenocarcinoma | Reference | Reference |
| SCC | 1.00 (0.79 - 1.26, 0.99) | * |
| Large Cell | 1.37 (0.80 - 2.33, 0.25) | * |
| Other | 1.09 (0.82 - 1.43, 0.56) | * |
| Laterality |  |  |
| Right | Reference | Reference |
| Left | 0.86 (0.69 - 1.05, 0.14) | * |
| Unpaired | - | * |
| Unknown | 1.15 (0.47 - 2.79, 0.75) | * |
| Tumor Location |  |  |
| Main bronchus | Reference | Reference |
| Upper lobe | 1.31 (0.77 - 2.21, 0.31) | 1.31 (0.77 - 2.22, 0.32) |
| Middle lobe | 2.06 (1.04 - 4.04, 0.04) | 2.00 (1.01 - 3.95, 0.05) |
| Lower lobe | 1.99 (1.16 - 3.39, 0.01) | 1.97 (1.15 - 3.39, 0.01) |
| Lung NOS | 1.42 (0.72 - 2.79, 0.31) | 1.32 (0.67 - 2.61, 0.42) |
| Other | 2.81 (1.02 - 7.74, 0.05) | 3.13 (1.13 - 8.67, 0.03) |
| PET |  |  |
| No | Reference | Reference |
| Yes | 1.89 (1.13 - 3.17, 0.02) | 1.85 (1.10 - 3.11, 0.02) |
| # of Positive Nodes |  |  |
| 0 | Reference | Reference |
| 1-3 | 1.34 (0.74 - 2.43, 0.33) | * |
| 4+ | 1.08 (0.48 - 2.43, 0.85) | * |
| Unknown | 1.35 (0.79 - 2.30, 0.27) | * |
| Treatment Type |  |  |
| Trimodality | Reference | Reference |
| Chemotherapy & radiation | 1.97 (1.22 - 3.16, <.01) | 2.24 (1.36 - 3.69, <.01) |
| Surgery & radiation | 1.86 (0.78 - 4.45, 0.16) | 2.03 (0.84 - 4.91, 0.12) |
| Radiation alone | 1.75 (1.03 - 2.99, 0.04) | 1.95 (1.10 - 3.44, 0.02) |
| # of RT Fractions |  |  |
| 25 - 29 | Reference | Reference |
| 30 - 34 | 1.03 (0.78 - 1.37, 0.82) | 0.91 (0.68 - 1.21, 0.50) |
| 35 - 40 | 0.98 (0.73 - 1.30, 0.87) | 0.83 (0.62 - 1.13, 0.23) |
| Type of Treatment Center |  |  |
| Free Standing | Reference | Reference |
| Hospital Based | 1.31 (1.05 - 1.64, 0.02) | * |
| Both | 1.61 (0.51 - 5.06, 0.42) | * |
| Rural vs. Urban |  |  |
| Rural | Reference | Reference |
| Urban | 1.07 (0.82 - 1.39, 0.62) | * |
| Radiation Oncologist Density |  |  |
| 1st quartile | Reference | Reference |
| 2nd quartile | 1.09 (0.84 - 1.43, 0.52) | * |
| 3rd quartile | 1.20 (0.91 - 1.58, 0.19) | * |
| 4th quartile | 1.11 (0.82 - 1.50, 0.52) | * |
| Unknown | 1.10 (0.41 - 2.99, 0.85) | * |
| General Surgeon Density |  |  |
| 1st quartile | Reference | Reference |
| 2nd quartile | 1.08 (0.81 - 1.45, 0.59) | * |
| 3rd quartile | 1.35 (1.03 - 1.78, 0.03) | * |
| 4th quartile | 1.35 (1.01 - 1.79, 0.04) | * |
| Unknown | 1.19 (0.44 - 3.24, 0.73) | * |
| Physician Experience |  |  |
| 1st quartile | Reference | Reference |
| 2nd quartile | 1.11 (0.83 - 1.48, 0.48) | * |
| 3rd quartile | 1.07 (0.80 - 1.43, 0.65) | * |
| 4th quartile | 1.07 (0.80 - 1.43, 0.64) | * |
| State |  |  |
| California | Reference | Reference |
| Connecticut | 1.33 (0.88 - 2.02, 0.17) | 1.31 (0.86 - 1.98, 0.21) |
| Georgia | 0.96 (0.67 - 1.38, 0.81) | 0.98 (0.68 - 1.42, 0.92) |
| Hawaii | 1.14 (0.42 - 3.12, 0.80) | 1.17 (0.43 - 3.20, 0.76) |
| Iowa | 0.90 (0.56 - 1.47, 0.68) | 0.89 (0.55 - 1.45, 0.64) |
| Kentucky | 0.72 (0.46 - 1.12, 0.14) | 0.72 (0.46 - 1.13, 0.16) |
| Louisiana | 1.32 (0.88 - 1.98, 0.18) | 1.30 (0.86 - 1.96, 0.21) |
| Michigan | 1.17 (0.77 - 1.78, 0.46) | 1.17 (0.77 - 1.78, 0.46) |
| New Jersey | 1.49 (1.07 - 2.07, 0.02) | 1.49 (1.07 - 2.08, 0.02) |
| New Mexico | 0.74 (0.27 - 2.01, 0.55) | 0.76 (0.28 - 2.07, 0.59) |
| Utah | 1.41 (0.52 - 3.86, 0.50) | 1.48 (0.54 - 4.08, 0.45) |
| Washington | 1.71 (1.15 - 2.56, <.01) | 1.73 (1.15 - 2.59, <.01) |
| Year of Diagnosis |  |  |
| 2006 | Reference | Reference |
| 2007 | 1.04 (0.76 - 1.44, 0.79) | * |
| 2008 | 0.94 (0.67 - 1.31, 0.70) | * |
| 2009 | 1.10 (0.79 - 1.52, 0.57) | * |
| 2010 | 0.60 (0.40 - 0.89, 0.01) | * |
| 2011 | 0.94 (0.67 - 1.33, 0.75) | * |
| IMRT |  |  |
| No | Reference | Reference |
| Yes | 1.11 (0.88 - 1.40, 0.38) | * |
| ^X^ Multivariate Cox regressions were performed using stepwise forward and backwards elimination with threshold values of p ≤ 0.20 and p ≤ 0.05, respectively.  * Covariate auto-excluded from model during forward or backward selection.  Abbrev: HR, hazard ratio. CI, confidence interval. | | |

| **Supplementary Table T7**. Multivariate proportional hazards regression for esophagus toxicity (all components).^X^ | | |
| --- | --- | --- |
| Parameter | Univariate  HR (95% CI, P-value) | Multivariate  HR (95% CI, P-Value) |
| Daily IGRT |  |  |
| No | Reference | Reference |
| Yes | 1.12 (1.00 - 1.24, 0.04) | 1.05 (0.93 - 1.18, 0.44) |
| Age |  |  |
| 65 - 74 | Reference | Reference |
| 75 - 84 | 1.03 (0.94 - 1.13, 0.55) | 1.10 (1.00 - 1.21, 0.04) |
| 85+ | 0.65 (0.52 - 0.81, <.01) | 0.83 (0.66 - 1.04, 0.11) |
| Race |  |  |
| White | Reference | Reference |
| Black | 0.85 (0.72 - 1.00, 0.05) | * |
| Hispanic | 1.08 (0.66 - 1.76, 0.76) | * |
| Other | 0.96 (0.77 - 1.21, 0.74) | * |
| COPD |  |  |
| No | Reference | Reference |
| Yes | 1.06 (0.97 - 1.15, 0.23) | * |
| Charlson Score (no COPD) |  |  |
| 0 | Reference | Reference |
| 1-2 | 0.99 (0.90 - 1.09, 0.84) | * |
| > 2 | 1.03 (0.88 - 1.20, 0.71) | * |
| Supplemental O2 |  |  |
| No | Reference | Reference |
| Yes | 0.98 (0.88 - 1.08, 0.66) | 0.98 (0.88 - 1.09, 0.69) |
| Homebound |  |  |
| No | Reference | Reference |
| Yes | 1.13 (0.82 - 1.57, 0.46) | 1.26 (0.90 - 1.75, 0.17) |
| Stage |  |  |
| Stage IIIA | Reference | Reference |
| Stage IIIB | 1.17 (1.07 - 1.27, <.01) | 1.13 (1.03 - 1.24, <.01) |
| T-Stage |  |  |
| TX | Reference | Reference |
| T0 | 0.52 (0.23 - 1.19, 0.12) | * |
| T1 | 0.88 (0.69 - 1.12, 0.30) | * |
| T2 | 0.96 (0.77 - 1.20, 0.71) | * |
| T3 | 0.86 (0.67 - 1.12, 0.26) | * |
| T4 | 0.98 (0.79 - 1.22, 0.86) | * |
| Tumor Size |  |  |
| < 2.0 | Reference | Reference |
| 2.0-5.0 | 1.05 (0.88 - 1.27, 0.58) | * |
| > 5.0 | 1.07 (0.88 - 1.29, 0.52) | * |
| Unknown | 1.24 (1.01 - 1.53, 0.04) | * |
| Histology |  |  |
| Adenocarcinoma | Reference | Reference |
| SCC | 1.07 (0.97 - 1.19, 0.18) | * |
| Large Cell | 1.05 (0.81 - 1.36, 0.73) | * |
| Other | 1.12 (0.99 - 1.27, 0.07) | * |
| Laterality |  |  |
| Right | Reference | Reference |
| Left | 0.96 (0.88 - 1.05, 0.39) | * |
| Unpaired | 0.31 (0.04 - 2.23, 0.25) | * |
| Unknown | 1.17 (0.75 - 1.83, 0.48) | * |
| Tumor Location |  |  |
| Main bronchus | Reference | Reference |
| Upper lobe | 0.73 (0.62 - 0.88, <.01) | 0.77 (0.65 - 0.92, <.01) |
| Middle lobe | 0.57 (0.42 - 0.78, <.01) | 0.62 (0.45 - 0.85, <.01) |
| Lower lobe | 0.88 (0.73 - 1.05, 0.16) | 0.92 (0.76 - 1.11, 0.40) |
| Lung NOS | 0.91 (0.71 - 1.16, 0.44) | 0.88 (0.69 - 1.13, 0.33) |
| Other | 0.71 (0.40 - 1.25, 0.23) | 0.82 (0.46 - 1.45, 0.49) |
| PET |  |  |
| No | Reference | Reference |
| Yes | 1.13 (0.95 - 1.34, 0.17) | * |
| # of Positive Nodes |  |  |
| 0 | Reference | Reference |
| 1-3 | 1.08 (0.84 - 1.40, 0.55) | * |
| 4+ | 1.05 (0.74 - 1.48, 0.78) | * |
| Unknown | 1.20 (0.96 - 1.50, 0.11) | * |
| Treatment Type |  |  |
| Trimodality | Reference | Reference |
| Chemotherapy & radiation | 1.37 (1.14 - 1.63, <.01) | 1.61 (1.33 - 1.94, <.01) |
| Surgery & radiation | 0.53 (0.33 - 0.85, <.01) | 0.53 (0.33 - 0.85, <.01) |
| Radiation alone | 0.76 (0.61 - 0.94, 0.01) | 0.85 (0.68 - 1.07, 0.16) |
| # of RT Fractions |  |  |
| 25 - 29 | Reference | Reference |
| 30 - 34 | 0.74 (0.66 - 0.83, <.01) | 0.68 (0.60 - 0.77, <.01) |
| 35 - 40 | 0.67 (0.59 - 0.75, <.01) | 0.58 (0.51 - 0.66, <.01) |
| Type of Treatment Center |  |  |
| Free Standing | Reference | Reference |
| Hospital Based | 1.00 (0.91 - 1.10, 0.97) | * |
| Both | 1.16 (0.67 - 2.01, 0.59) | * |
| Rural vs. Urban |  |  |
| Rural | Reference | Reference |
| Urban | 0.98 (0.88 - 1.10, 0.72) | * |
| Radiation Oncologist Density |  |  |
| 1st quartile | Reference | Reference |
| 2nd quartile | 0.97 (0.87 - 1.09, 0.60) | 0.80 (0.69 - 0.94, <.01) |
| 3rd quartile | 0.84 (0.74 - 0.95, <.01) | 0.77 (0.64 - 0.92, <.01) |
| 4th quartile | 0.83 (0.73 - 0.95, <.01) | 0.66 (0.54 - 0.80, <.01) |
| Unknown | 0.64 (0.39 - 1.05, 0.08) | 0.55 (0.33 - 0.91, 0.02) |
| General Surgeon Density |  |  |
| 1st quartile | Reference | Reference |
| 2nd quartile | 1.13 (1.00 - 1.27, 0.05) | 1.26 (1.09 - 1.47, <.01) |
| 3rd quartile | 0.93 (0.82 - 1.05, 0.24) | 1.22 (1.01 - 1.47, 0.04) |
| 4th quartile | 1.00 (0.88 - 1.13, 0.97) | 1.32 (1.09 - 1.61, <.01) |
| Unknown | 0.70 (0.43 - 1.16, 0.16) | * |
| Physician Experience |  |  |
| 1st quartile | Reference | Reference |
| 2nd quartile | 0.94 (0.83 - 1.07, 0.33) | * |
| 3rd quartile | 0.95 (0.84 - 1.07, 0.41) | * |
| 4th quartile | 0.86 (0.75 - 0.97, 0.02) | * |
| State |  |  |
| California | Reference | Reference |
| Connecticut | 1.00 (0.83 - 1.21, 0.99) | 1.04 (0.84 - 1.29, 0.71) |
| Georgia | 0.78 (0.67 - 0.90, <.01) | 0.80 (0.68 - 0.94, <.01) |
| Hawaii | 0.60 (0.36 - 0.98, 0.04) | * |
| Iowa | 0.73 (0.59 - 0.89, <.01) | 0.78 (0.63 - 0.96, 0.02) |
| Kentucky | 0.84 (0.71 - 0.99, 0.04) | 0.86 (0.72 - 1.03, 0.10) |
| Louisiana | 0.96 (0.80 - 1.14, 0.61) | 0.86 (0.71 - 1.04, 0.13) |
| Michigan | 0.67 (0.55 - 0.82, <.01) | 0.67 (0.51 - 0.88, <.01) |
| New Jersey | 0.79 (0.68 - 0.92, <.01) | 0.77 (0.64 - 0.91, <.01) |
| New Mexico | 0.86 (0.60 - 1.23, 0.40) | 0.75 (0.52 - 1.08, 0.12) |
| Utah | 0.73 (0.44 - 1.23, 0.24) | 0.64 (0.38 - 1.07, 0.09) |
| Washington | 0.94 (0.77 - 1.15, 0.56) | 0.96 (0.76 - 1.20, 0.70) |
| Year of Diagnosis |  |  |
| 2006 | Reference | Reference |
| 2007 | 1.01 (0.87 - 1.16, 0.94) | * |
| 2008 | 0.89 (0.76 - 1.03, 0.12) | * |
| 2009 | 1.03 (0.88 - 1.19, 0.74) | * |
| 2010 | 0.99 (0.85 - 1.16, 0.93) | * |
| 2011 | 1.02 (0.87 - 1.19, 0.83) | * |
| IMRT |  |  |
| No | Reference | Reference |
| Yes | 1.22 (1.10 - 1.35, <.01) | 1.20 (1.06 - 1.34, <.01) |
| ^X^ Multivariate Cox regressions were performed using stepwise forward and backwards elimination with threshold values of p ≤ 0.20 and p ≤ 0.05, respectively.  * Covariate auto-excluded from model during forward or backward selection.  Abbrev: HR, hazard ratio. CI, confidence interval. | | |

| **Supplementary Table T8**. Multivariate proportional hazards regression for esophagitis (based on diagnosis codes).^X^ | | |
| --- | --- | --- |
| Parameter | Univariate  HR (95% CI, P-value) | Multivariate  HR (95% CI, P-Value) |
| Daily IGRT |  |  |
| No | Reference | Reference |
| Yes | 1.19 (1.03 - 1.37, 0.02) | 1.27 (1.08 - 1.49, <.01) |
| Age |  |  |
| 65 - 74 | Reference | Reference |
| 75 - 84 | 0.97 (0.85 - 1.10, 0.59) | 1.05 (0.92 - 1.20, 0.48) |
| 85+ | 0.59 (0.42 - 0.81, <.01) | 0.79 (0.56 - 1.11, 0.17) |
| Race |  |  |
| White | Reference | Reference |
| Black | 0.91 (0.73 - 1.15, 0.44) | * |
| Hispanic | 1.44 (0.79 - 2.60, 0.23) | * |
| Other | 0.88 (0.63 - 1.22, 0.45) | * |
| COPD |  |  |
| No | Reference | Reference |
| Yes | 1.20 (1.06 - 1.35, <.01) | 1.18 (1.04 - 1.36, 0.01) |
| Charlson Score (no COPD) |  |  |
| 0 | Reference | Reference |
| 1-2 | 1.02 (0.89 - 1.16, 0.79) | * |
| > 2 | 1.11 (0.90 - 1.37, 0.35) | * |
| Supplemental O2 |  |  |
| No | Reference | Reference |
| Yes | 1.07 (0.93 - 1.23, 0.34) | 0.98 (0.84 - 1.14, 0.78) |
| Homebound |  |  |
| No | Reference | Reference |
| Yes | 1.16 (0.73 - 1.84, 0.54) | 1.12 (0.70 - 1.81, 0.64) |
| Stage |  |  |
| Stage IIIA | Reference | Reference |
| Stage IIIB | 1.14 (1.00 - 1.29, 0.04) | 1.11 (0.98 - 1.26, 0.09) |
| T-Stage |  |  |
| TX | Reference | Reference |
| T0 | 1.01 (0.40 - 2.55, 0.98) | * |
| T1 | 0.96 (0.69 - 1.34, 0.81) | * |
| T2 | 0.95 (0.70 - 1.29, 0.73) | * |
| T3 | 0.91 (0.64 - 1.29, 0.60) | * |
| T4 | 0.96 (0.71 - 1.30, 0.80) | * |
| Tumor Size |  |  |
| < 2.0 | Reference | Reference |
| 2.0-5.0 | 1.04 (0.80 - 1.34, 0.77) | * |
| > 5.0 | 0.94 (0.72 - 1.22, 0.64) | * |
| Unknown | 1.25 (0.95 - 1.66, 0.11) | * |
| Histology |  |  |
| Adenocarcinoma | Reference | Reference |
| SCC | 1.13 (0.97 - 1.30, 0.11) | * |
| Large Cell | 1.20 (0.84 - 1.70, 0.32) | * |
| Other | 1.12 (0.94 - 1.33, 0.20) | * |
| Laterality |  |  |
| Right | Reference | Reference |
| Left | 1.04 (0.92 - 1.18, 0.52) | * |
| Unpaired | 0.76 (0.11 - 5.37, 0.78) | * |
| Unknown | 1.84 (1.12 - 3.03, 0.02) | * |
| Tumor Location |  |  |
| Main bronchus | Reference | Reference |
| Upper lobe | 0.78 (0.61 - 0.99, 0.05) | * |
| Middle lobe | 0.56 (0.36 - 0.87, 0.01) | * |
| Lower lobe | 0.81 (0.63 - 1.06, 0.13) | * |
| Lung NOS | 1.09 (0.78 - 1.52, 0.61) | * |
| Other | 0.76 (0.35 - 1.66, 0.50) | * |
| PET |  |  |
| No | Reference | Reference |
| Yes | 1.19 (0.92 - 1.52, 0.18) | * |
| # of Positive Nodes |  |  |
| 0 | Reference | Reference |
| 1-3 | 1.23 (0.87 - 1.74, 0.24) | * |
| 4+ | 0.72 (0.42 - 1.22, 0.22) | * |
| Unknown | 1.17 (0.86 - 1.59, 0.33) | * |
| Treatment Type |  |  |
| Trimodality | Reference | Reference |
| Chemotherapy & radiation | 1.39 (1.08 - 1.77, <.01) | 1.73 (1.33 - 2.24, <.01) |
| Surgery & radiation | 0.43 (0.20 - 0.94, 0.03) | 0.44 (0.20 - 0.97, 0.04) |
| Radiation alone | 0.81 (0.60 - 1.09, 0.17) | 0.97 (0.70 - 1.34, 0.86) |
| # of RT Fractions |  |  |
| 25 - 29 | Reference | Reference |
| 30 - 34 | 0.70 (0.60 - 0.82, <.01) | 0.64 (0.54 - 0.75, <.01) |
| 35 - 40 | 0.63 (0.54 - 0.74, <.01) | 0.53 (0.45 - 0.63, <.01) |
| Type of Treatment Center |  |  |
| Free Standing | Reference | Reference |
| Hospital Based | 1.04 (0.91 - 1.18, 0.61) | * |
| Both | 0.81 (0.33 - 1.96, 0.64) | * |
| Rural vs. Urban |  |  |
| Rural | Reference | Reference |
| Urban | 0.81 (0.70 - 0.94, <.01) | * |
| Radiation Oncologist Density |  |  |
| 1st quartile | Reference | Reference |
| 2nd quartile | 0.83 (0.71 - 0.97, 0.02) | * |
| 3rd quartile | 0.70 (0.58 - 0.83, <.01) | * |
| 4th quartile | 0.73 (0.61 - 0.89, <.01) | * |
| Unknown | 0.59 (0.29 - 1.19, 0.14) | * |
| General Surgeon Density |  |  |
| 1st quartile | Reference | Reference |
| 2nd quartile | 1.00 (0.85 - 1.18, 1.00) | * |
| 3rd quartile | 0.72 (0.61 - 0.86, <.01) | * |
| 4th quartile | 0.89 (0.75 - 1.06, 0.20) | * |
| Unknown | 0.64 (0.32 - 1.30, 0.22) | * |
| Physician Experience |  |  |
| 1st quartile | Reference | Reference |
| 2nd quartile | 1.03 (0.87 - 1.23, 0.73) | * |
| 3rd quartile | 1.08 (0.91 - 1.29, 0.38) | * |
| 4th quartile | 0.97 (0.81 - 1.16, 0.72) | * |
| State |  |  |
| California | Reference | Reference |
| Connecticut | 0.79 (0.59 - 1.05, 0.10) | 0.79 (0.59 - 1.05, 0.10) |
| Georgia | 1.04 (0.85 - 1.27, 0.71) | 1.03 (0.84 - 1.27, 0.74) |
| Hawaii | 0.68 (0.33 - 1.37, 0.28) | 0.66 (0.32 - 1.33, 0.24) |
| Iowa | 0.79 (0.59 - 1.05, 0.10) | 0.80 (0.60 - 1.07, 0.14) |
| Kentucky | 1.05 (0.84 - 1.32, 0.65) | 1.05 (0.84 - 1.31, 0.68) |
| Louisiana | 1.46 (1.17 - 1.83, <.01) | 1.55 (1.23 - 1.94, <.01) |
| Michigan | 0.57 (0.42 - 0.78, <.01) | 0.55 (0.41 - 0.75, <.01) |
| New Jersey | 0.97 (0.79 - 1.20, 0.80) | 0.95 (0.77 - 1.18, 0.66) |
| New Mexico | 0.89 (0.53 - 1.50, 0.65) | 0.89 (0.53 - 1.50, 0.66) |
| Utah | 0.63 (0.28 - 1.41, 0.26) | 0.58 (0.25 - 1.30, 0.18) |
| Washington | 0.74 (0.54 - 1.01, 0.05) | 0.74 (0.54 - 1.01, 0.06) |
| Year of Diagnosis |  |  |
| 2006 | Reference | Reference |
| 2007 | 0.96 (0.79 - 1.17, 0.67) | 0.94 (0.77 - 1.15, 0.55) |
| 2008 | 0.75 (0.61 - 0.93, <.01) | 0.73 (0.59 - 0.90, <.01) |
| 2009 | 0.83 (0.67 - 1.03, 0.09) | 0.75 (0.61 - 0.94, 0.01) |
| 2010 | 0.88 (0.71 - 1.09, 0.26) | 0.80 (0.64 - 1.00, 0.05) |
| 2011 | 1.02 (0.83 - 1.25, 0.85) | 0.87 (0.70 - 1.09, 0.22) |
| IMRT |  |  |
| No | Reference | Reference |
| Yes | 1.19 (1.03 - 1.37, 0.02) | * |
| ^X^ Multivariate Cox regressions were performed using stepwise forward and backwards elimination with threshold values of p ≤ 0.20 and p ≤ 0.05, respectively.  * Covariate auto-excluded from model during forward or backward selection.  Abbrev: HR, hazard ratio. CI, confidence interval. | | |

| **Supplementary Table T9**. Multivariate proportional hazards regression for dehydration (based on diagnosis codes).^X^ | | |
| --- | --- | --- |
| Parameter | Univariate  HR (95% CI, P-value) | Multivariate  HR (95% CI, P-Value) |
| Daily IGRT |  |  |
| No | Reference | Reference |
| Yes | 1.13 (1.00 - 1.28, 0.04) | 1.06 (0.92 - 1.21, 0.41) |
| Age |  |  |
| 65 - 74 | Reference | Reference |
| 75 - 84 | 1.10 (0.99 - 1.22, 0.07) | 1.19 (1.07 - 1.32, <.01) |
| 85+ | 0.68 (0.53 - 0.88, <.01) | 0.92 (0.71 - 1.19, 0.52) |
| Race |  |  |
| White | Reference | Reference |
| Black | 0.72 (0.59 - 0.87, <.01) | 0.76 (0.62 - 0.93, <.01) |
| Hispanic | 0.78 (0.42 - 1.46, 0.44) | 0.74 (0.40 - 1.38, 0.34) |
| Other | 1.00 (0.78 - 1.29, 0.98) | 0.94 (0.72 - 1.24, 0.67) |
| COPD |  |  |
| No | Reference | Reference |
| Yes | 1.02 (0.93 - 1.13, 0.64) | * |
| Charlson Score (no COPD) |  |  |
| 0 | Reference | Reference |
| 1-2 | 0.96 (0.86 - 1.08, 0.51) | * |
| > 2 | 0.90 (0.75 - 1.08, 0.28) | * |
| Supplemental O2 |  |  |
| No | Reference | Reference |
| Yes | 0.98 (0.87 - 1.10, 0.73) | 0.99 (0.88 - 1.11, 0.85) |
| Homebound |  |  |
| No | Reference | Reference |
| Yes | 1.10 (0.76 - 1.60, 0.62) | 1.24 (0.85 - 1.82, 0.27) |
| Stage |  |  |
| Stage IIIA | Reference | Reference |
| Stage IIIB | 1.18 (1.07 - 1.31, <.01) | 1.13 (1.02 - 1.25, 0.02) |
| T-Stage |  |  |
| TX | Reference | Reference |
| T0 | 0.39 (0.14 - 1.06, 0.07) | * |
| T1 | 0.75 (0.58 - 0.98, 0.04) | * |
| T2 | 0.88 (0.69 - 1.12, 0.29) | * |
| T3 | 0.77 (0.58 - 1.02, 0.06) | * |
| T4 | 0.90 (0.70 - 1.14, 0.37) | * |
| Tumor Size |  |  |
| < 2.0 | Reference | Reference |
| 2.0-5.0 | 1.13 (0.91 - 1.40, 0.26) | * |
| > 5.0 | 1.15 (0.92 - 1.44, 0.21) | * |
| Unknown | 1.32 (1.04 - 1.67, 0.02) | * |
| Histology |  |  |
| Adenocarcinoma | Reference | Reference |
| SCC | 1.05 (0.93 - 1.18, 0.43) | * |
| Large Cell | 1.03 (0.76 - 1.39, 0.84) | * |
| Other | 1.14 (0.99 - 1.31, 0.07) | * |
| Laterality |  |  |
| Right | Reference | Reference |
| Left | 0.92 (0.83 - 1.02, 0.12) | * |
| Unpaired | 0.00 (0.00 - 4E78, 0.92) | * |
| Unknown | 0.84 (0.49 - 1.46, 0.54) | * |
| Tumor Location |  |  |
| Main bronchus | Reference | Reference |
| Upper lobe | 0.76 (0.62 - 0.94, <.01) | 0.81 (0.66 - 1.00, 0.05) |
| Middle lobe | 0.64 (0.45 - 0.91, 0.01) | 0.72 (0.50 - 1.02, 0.07) |
| Lower lobe | 1.00 (0.81 - 1.24, 0.99) | 1.06 (0.85 - 1.31, 0.61) |
| Lung NOS | 1.02 (0.77 - 1.34, 0.92) | 0.99 (0.75 - 1.32, 0.97) |
| Other | 0.94 (0.52 - 1.71, 0.84) | 1.13 (0.62 - 2.05, 0.70) |
| PET |  |  |
| No | Reference | Reference |
| Yes | 1.12 (0.92 - 1.36, 0.26) | * |
| # of Positive Nodes |  |  |
| 0 | Reference | Reference |
| 1-3 | 0.99 (0.74 - 1.33, 0.97) | * |
| 4+ | 1.06 (0.71 - 1.56, 0.78) | * |
| Unknown | 1.21 (0.94 - 1.57, 0.14) | * |
| Treatment Type |  |  |
| Trimodality | Reference | Reference |
| Chemotherapy & radiation | 1.51 (1.23 - 1.86, <.01) | 1.80 (1.44 - 2.24, <.01) |
| Surgery & radiation | 0.55 (0.31 - 0.96, 0.04) | 0.53 (0.30 - 0.94, 0.03) |
| Radiation alone | 0.72 (0.55 - 0.93, 0.01) | 0.79 (0.60 - 1.05, 0.10) |
| # of RT Fractions |  |  |
| 25 - 29 | Reference | Reference |
| 30 - 34 | 0.75 (0.66 - 0.85, <.01) | 0.67 (0.59 - 0.77, <.01) |
| 35 - 40 | 0.65 (0.56 - 0.74, <.01) | 0.55 (0.48 - 0.64, <.01) |
| Type of Treatment Center |  |  |
| Free Standing | Reference | Reference |
| Hospital Based | 0.96 (0.87 - 1.07, 0.48) | * |
| Both | 1.21 (0.67 - 2.20, 0.53) | * |
| Rural vs. Urban |  |  |
| Rural | Reference | Reference |
| Urban | 1.06 (0.93 - 1.21, 0.34) | * |
| Radiation Oncologist Density |  |  |
| 1st quartile | Reference | Reference |
| 2nd quartile | 1.05 (0.92 - 1.19, 0.50) | 0.86 (0.72 - 1.03, 0.09) |
| 3rd quartile | 0.91 (0.79 - 1.05, 0.18) | 0.80 (0.66 - 0.98, 0.03) |
| 4th quartile | 0.84 (0.72 - 0.98, 0.03) | 0.66 (0.53 - 0.83, <.01) |
| Unknown | 0.74 (0.43 - 1.29, 0.29) | 0.63 (0.35 - 1.13, 0.12) |
| General Surgeon Density |  |  |
| 1st quartile | Reference | Reference |
| 2nd quartile | 1.17 (1.02 - 1.34, 0.03) | 1.27 (1.07 - 1.51, <.01) |
| 3rd quartile | 0.99 (0.86 - 1.13, 0.83) | 1.22 (0.99 - 1.52, 0.06) |
| 4th quartile | 1.00 (0.87 - 1.16, 0.96) | 1.33 (1.06 - 1.66, 0.01) |
| Unknown | 0.80 (0.46 - 1.39, 0.43) | * |
| Physician Experience |  |  |
| 1st quartile | Reference | Reference |
| 2nd quartile | 0.93 (0.81 - 1.07, 0.31) | * |
| 3rd quartile | 0.88 (0.77 - 1.02, 0.08) | * |
| 4th quartile | 0.80 (0.69 - 0.92, <.01) | * |
| State |  |  |
| California | Reference | Reference |
| Connecticut | 1.07 (0.87 - 1.31, 0.53) | 1.09 (0.86 - 1.38, 0.47) |
| Georgia | 0.74 (0.62 - 0.87, <.01) | 0.77 (0.65 - 0.93, <.01) |
| Hawaii | 0.65 (0.37 - 1.12, 0.12) | * |
| Iowa | 0.70 (0.56 - 0.88, <.01) | 0.75 (0.59 - 0.95, 0.02) |
| Kentucky | 0.78 (0.65 - 0.94, <.01) | 0.81 (0.66 - 1.00, 0.04) |
| Louisiana | 0.83 (0.68 - 1.02, 0.07) | 0.78 (0.63 - 0.98, 0.03) |
| Michigan | 0.71 (0.57 - 0.88, <.01) | 0.71 (0.52 - 0.97, 0.03) |
| New Jersey | 0.72 (0.61 - 0.86, <.01) | 0.68 (0.56 - 0.83, <.01) |
| New Mexico | 0.88 (0.59 - 1.31, 0.52) | 0.79 (0.53 - 1.19, 0.27) |
| Utah | 0.84 (0.48 - 1.46, 0.54) | 0.72 (0.41 - 1.27, 0.26) |
| Washington | 1.01 (0.81 - 1.25, 0.94) | 1.01 (0.78 - 1.30, 0.93) |
| Year of Diagnosis |  |  |
| 2006 | Reference | Reference |
| 2007 | 1.07 (0.90 - 1.26, 0.45) | * |
| 2008 | 0.91 (0.77 - 1.09, 0.31) | * |
| 2009 | 1.09 (0.92 - 1.30, 0.29) | * |
| 2010 | 1.03 (0.87 - 1.23, 0.71) | * |
| 2011 | 1.05 (0.88 - 1.26, 0.57) | * |
| IMRT |  |  |
| No | Reference | Reference |
| Yes | 1.22 (1.08 - 1.37, <.01) | 1.18 (1.03 - 1.35, 0.01) |
| ^X^ Multivariate Cox regressions were performed using stepwise forward and backwards elimination with threshold values of p ≤ 0.20 and p ≤ 0.05, respectively.  * Covariate auto-excluded from model during forward or backward selection.  Abbrev: HR, hazard ratio. CI, confidence interval. | | |

| **Supplementary Table T10**. Multivariate proportional hazards regression for feeding tube placement (based on procedural codes).^X^ | | |
| --- | --- | --- |
| Parameter | Univariate  HR (95% CI, P-value) | Multivariate  HR (95% CI, P-Value) |
| Daily IGRT |  |  |
| No | Reference | Reference |
| Yes | 1.16 (0.81 - 1.65, 0.43) | 1.07 (0.74 - 1.54, 0.72) |
| Age |  |  |
| 65 - 74 | Reference | Reference |
| 75 - 84 | 0.86 (0.64 - 1.16, 0.34) | 0.91 (0.67 - 1.24, 0.54) |
| 85+ | 0.36 (0.11 - 1.13, 0.08) | 0.46 (0.14 - 1.49, 0.19) |
| Race |  |  |
| White | Reference | Reference |
| Black | 0.91 (0.56 - 1.50, 0.72) | * |
| Hispanic | 0.99 (0.14 - 7.06, 0.99) | * |
| Other | 1.10 (0.58 - 2.09, 0.77) | * |
| COPD |  |  |
| No | Reference | Reference |
| Yes | 1.18 (0.89 - 1.56, 0.26) | * |
| Charlson Score (no COPD) |  |  |
| 0 | Reference | Reference |
| 1-2 | 0.83 (0.61 - 1.12, 0.22) | * |
| > 2 | 0.69 (0.38 - 1.24, 0.22) | * |
| Supplemental O2 |  |  |
| No | Reference | Reference |
| Yes | 1.02 (0.72 - 1.46, 0.89) | 0.97 (0.67 - 1.40, 0.87) |
| Homebound |  |  |
| No | Reference | Reference |
| Yes | 0.47 (0.12 - 1.90, 0.29) | 0.29 (0.07 - 1.26, 0.10) |
| Stage |  |  |
| Stage IIIA | Reference | Reference |
| Stage IIIB | 1.46 (1.10 - 1.95, <.01) | 1.44 (1.07 - 1.94, 0.02) |
| T-Stage |  |  |
| TX | Reference | Reference |
| T0 | 1.62 (0.44 - 6.02, 0.47) | * |
| T1 | 1.35 (0.64 - 2.86, 0.43) | * |
| T2 | 1.07 (0.52 - 2.18, 0.86) | * |
| T3 | 0.98 (0.44 - 2.17, 0.95) | * |
| T4 | 1.49 (0.75 - 2.98, 0.26) | * |
| Tumor Size |  |  |
| < 2.0 | Reference | Reference |
| 2.0-5.0 | 1.17 (0.63 - 2.18, 0.62) | * |
| > 5.0 | 0.95 (0.49 - 1.82, 0.87) | * |
| Unknown | 1.19 (0.60 - 2.36, 0.61) | * |
| Histology |  |  |
| Adenocarcinoma | Reference | Reference |
| SCC | 1.31 (0.94 - 1.84, 0.11) | * |
| Large Cell | 1.89 (0.81 - 4.43, 0.14) | * |
| Other | 1.22 (0.81 - 1.85, 0.34) | * |
| Laterality |  |  |
| Right | Reference | Reference |
| Left | 1.05 (0.79 - 1.40, 0.74) | * |
| Unpaired | - | * |
| Unknown | 0.67 (0.16 - 2.70, 0.57) | * |
| Tumor Location |  |  |
| Main bronchus | Reference | Reference |
| Upper lobe | 0.51 (0.29 - 0.88, 0.02) | 0.60 (0.34 - 1.06, 0.08) |
| Middle lobe | 0.53 (0.21 - 1.37, 0.19) | 0.77 (0.29 - 2.01, 0.59) |
| Lower lobe | 0.81 (0.46 - 1.43, 0.46) | 1.02 (0.57 - 1.83, 0.94) |
| Lung NOS | 0.55 (0.27 - 1.13, 0.10) | 0.61 (0.29 - 1.28, 0.19) |
| Other | 0.66 (0.09 - 5.00, 0.69) | 0.77 (0.10 - 5.92, 0.81) |
| PET |  |  |
| No | Reference | Reference |
| Yes | 1.14 (0.66 - 1.97, 0.63) | * |
| # of Positive Nodes |  |  |
| 0 | Reference | Reference |
| 1-3 | 1.08 (0.49 - 2.40, 0.85) | * |
| 4+ | 0.95 (0.34 - 2.64, 0.93) | * |
| Unknown | 1.11 (0.54 - 2.26, 0.77) | * |
| Treatment Type |  |  |
| Trimodality | Reference | Reference |
| Chemotherapy & radiation | 1.05 (0.65 - 1.69, 0.85) | 1.19 (0.70 - 2.01, 0.52) |
| Surgery & radiation | 0.35 (0.05 - 2.62, 0.31) | 0.38 (0.05 - 2.86, 0.35) |
| Radiation alone | 0.64 (0.34 - 1.23, 0.18) | 0.77 (0.38 - 1.58, 0.48) |
| # of RT Fractions |  |  |
| 25 - 29 | Reference | Reference |
| 30 - 34 | 0.63 (0.45 - 0.89, <.01) | 0.55 (0.38 - 0.79, <.01) |
| 35 - 40 | 0.50 (0.34 - 0.72, <.01) | 0.42 (0.29 - 0.63, <.01) |
| Type of Treatment Center |  |  |
| Free Standing | Reference | Reference |
| Hospital Based | 0.88 (0.66 - 1.19, 0.41) | * |
| Both | 1.19 (0.16 - 8.56, 0.87) | * |
| Rural vs. Urban |  |  |
| Rural | Reference | Reference |
| Urban | 0.76 (0.54 - 1.08, 0.13) | * |
| Radiation Oncologist Density |  |  |
| 1st quartile | Reference | Reference |
| 2nd quartile | 0.47 (0.32 - 0.69, <.01) | 0.47 (0.32 - 0.69, <.01) |
| 3rd quartile | 0.80 (0.56 - 1.15, 0.23) | 0.86 (0.60 - 1.25, 0.44) |
| 4th quartile | 0.43 (0.27 - 0.69, <.01) | 0.38 (0.24 - 0.61, <.01) |
| Unknown | 0.53 (0.07 - 3.83, 0.53) | 0.48 (0.07 - 3.44, 0.46) |
| General Surgeon Density |  |  |
| 1st quartile | Reference | Reference |
| 2nd quartile | 0.81 (0.57 - 1.16, 0.25) | * |
| 3rd quartile | 0.53 (0.36 - 0.78, <.01) | * |
| 4th quartile | 0.46 (0.30 - 0.72, <.01) | * |
| Unknown | 0.54 (0.08 - 3.91, 0.54) | * |
| Physician Experience |  |  |
| 1st quartile | Reference | Reference |
| 2nd quartile | 1.12 (0.75 - 1.69, 0.58) | * |
| 3rd quartile | 1.02 (0.69 - 1.51, 0.92) | * |
| 4th quartile | 1.25 (0.84 - 1.84, 0.27) | * |
| State |  |  |
| California | Reference | Reference |
| Connecticut | 0.52 (0.24 - 1.11, 0.09) | * |
| Georgia | 1.14 (0.72 - 1.83, 0.57) | * |
| Hawaii | 0.78 (0.11 - 5.70, 0.81) | * |
| Iowa | 0.78 (0.39 - 1.57, 0.49) | * |
| Kentucky | 1.39 (0.86 - 2.23, 0.17) | * |
| Louisiana | 1.73 (1.05 - 2.85, 0.03) | * |
| Michigan | 0.94 (0.52 - 1.70, 0.84) | * |
| New Jersey | 0.95 (0.57 - 1.56, 0.83) | * |
| New Mexico | 1.37 (0.49 - 3.84, 0.55) | * |
| Utah | 1.98 (0.48 - 8.21, 0.35) | * |
| Washington | 0.53 (0.21 - 1.35, 0.18) | * |
| Year of Diagnosis |  |  |
| 2006 | Reference | Reference |
| 2007 | 1.13 (0.72 - 1.76, 0.60) | * |
| 2008 | 0.96 (0.60 - 1.53, 0.85) | * |
| 2009 | 1.14 (0.71 - 1.84, 0.59) | * |
| 2010 | 1.07 (0.64 - 1.79, 0.80) | * |
| 2011 | 1.30 (0.78 - 2.16, 0.32) | * |
| IMRT |  |  |
| No | Reference | Reference |
| Yes | 1.08 (0.78 - 1.49, 0.65) | * |
| ^X^ Multivariate Cox regressions were performed using stepwise forward and backwards elimination with threshold values of p ≤ 0.20 and p ≤ 0.05, respectively.  * Covariate auto-excluded from model during forward or backward selection.  Abbrev: HR, hazard ratio. CI, confidence interval. | | |

| **Supplementary Table T11**. Multivariate proportional hazards regression for mucositis (based on diagnosis codes).^X^ | | |
| --- | --- | --- |
| Parameter | Univariate  HR (95% CI, P-value) | Multivariate  HR (95% CI, P-Value) |
| Daily IGRT |  |  |
| No | Reference | Reference |
| Yes | 1.70 (1.03 - 2.78, 0.04) | 1.70 (1.03 - 2.80, 0.04) |
| Age |  |  |
| 65 - 74 | Reference | Reference |
| 75 - 84 | 0.72 (0.44 - 1.19, 0.20) | 0.93 (0.56 - 1.54, 0.77) |
| 85+ | 0.84 (0.30 - 2.34, 0.74) | 1.93 (0.68 - 5.44, 0.22) |
| Race |  |  |
| White | Reference | Reference |
| Black | 0.77 (0.31 - 1.92, 0.58) | * |
| Hispanic | 1.73 (0.24 - 12.5, 0.59) | * |
| Other | 0.65 (0.16 - 2.67, 0.55) | * |
| COPD |  |  |
| No | Reference | Reference |
| Yes | 1.24 (0.78 - 1.97, 0.36) | * |
| Charlson Score (no COPD) |  |  |
| 0 | Reference | Reference |
| 1-2 | 1.10 (0.68 - 1.80, 0.70) | * |
| > 2 | 0.94 (0.40 - 2.21, 0.88) | * |
| Supplemental O2 |  |  |
| No | Reference | Reference |
| Yes | 1.36 (0.82 - 2.25, 0.23) | 1.45 (0.87 - 2.40, 0.15) |
| Homebound |  |  |
| No | Reference | Reference |
| Yes | - | 0.00 (0.00 - . , 0.98) |
| Stage |  |  |
| Stage IIIA | Reference | Reference |
| Stage IIIB | 1.40 (0.88 - 2.24, 0.16) | 1.41 (0.87 - 2.26, 0.16) |
| T-Stage |  |  |
| TX | Reference | Reference |
| T0 | - | * |
| T1 | 0.84 (0.22 - 3.18, 0.80) | * |
| T2 | 1.01 (0.30 - 3.38, 0.98) | * |
| T3 | 1.23 (0.33 - 4.65, 0.76) | * |
| T4 | 1.15 (0.35 - 3.75, 0.82) | * |
| Tumor Size |  |  |
| < 2.0 | Reference | Reference |
| 2.0-5.0 | 0.65 (0.29 - 1.49, 0.31) | * |
| > 5.0 | 0.78 (0.34 - 1.81, 0.56) | * |
| Unknown | 0.67 (0.26 - 1.77, 0.42) | * |
| Histology |  |  |
| Adenocarcinoma | Reference | Reference |
| SCC | 1.04 (0.61 - 1.76, 0.90) | * |
| Large Cell | 1.37 (0.41 - 4.54, 0.61) | * |
| Other | 0.90 (0.46 - 1.73, 0.74) | * |
| Laterality |  |  |
| Right | Reference | Reference |
| Left | 1.15 (0.72 - 1.83, 0.56) | * |
| Unpaired | - | * |
| Unknown | - | * |
| Tumor Location |  |  |
| Main bronchus | Reference | Reference |
| Upper lobe | 0.88 (0.35 - 2.21, 0.78) | * |
| Middle lobe | 0.32 (0.04 - 2.74, 0.30) | * |
| Lower lobe | 1.01 (0.38 - 2.69, 0.98) | * |
| Lung NOS | 0.23 (0.03 - 1.93, 0.17) | * |
| Other | 1.65 (0.19 - 14.1, 0.65) | * |
| PET |  |  |
| No | Reference | Reference |
| Yes | 1.83 (0.58 - 5.81, 0.31) | * |
| # of Positive Nodes |  |  |
| 0 | Reference | Reference |
| 1-3 | 1.11 (0.30 - 4.10, 0.87) | * |
| 4+ | 1.54 (0.31 - 7.63, 0.60) | * |
| Unknown | 1.14 (0.36 - 3.63, 0.83) | * |
| Treatment Type |  |  |
| Trimodality | Reference | Reference |
| Chemotherapy & radiation | 1.03 (0.47 - 2.25, 0.94) | 1.08 (0.47 - 2.47, 0.86) |
| Surgery & radiation | - | 0.00 (0.00 - . , 0.98) |
| Radiation alone | 0.08 (0.01 - 0.61, 0.02) | 0.07 (0.01 - 0.58, 0.01) |
| # of RT Fractions |  |  |
| 25 - 29 | Reference | Reference |
| 30 - 34 | 0.64 (0.36 - 1.13, 0.12) | 0.64 (0.36 - 1.15, 0.14) |
| 35 - 40 | 0.58 (0.32 - 1.06, 0.08) | 0.52 (0.28 - 0.98, 0.04) |
| Type of Treatment Center |  |  |
| Free Standing | Reference | Reference |
| Hospital Based | 1.09 (0.67 - 1.79, 0.73) | * |
| Both | - | * |
| Rural vs. Urban |  |  |
| Rural | Reference | Reference |
| Urban | 0.49 (0.30 - 0.81, <.01) | 0.49 (0.30 - 0.81, <.01) |
| Radiation Oncologist Density |  |  |
| 1st quartile | Reference | Reference |
| 2nd quartile | 0.79 (0.45 - 1.39, 0.41) | * |
| 3rd quartile | 0.69 (0.37 - 1.29, 0.25) | * |
| 4th quartile | 0.43 (0.19 - 0.97, 0.04) | * |
| Unknown | - | * |
| General Surgeon Density |  |  |
| 1st quartile | Reference | Reference |
| 2nd quartile | 0.93 (0.52 - 1.65, 0.81) | * |
| 3rd quartile | 0.58 (0.30 - 1.11, 0.10) | * |
| 4th quartile | 0.57 (0.29 - 1.15, 0.12) | * |
| Unknown | - | * |
| Physician Experience |  |  |
| 1st quartile | Reference | Reference |
| 2nd quartile | 0.84 (0.43 - 1.64, 0.61) | * |
| 3rd quartile | 1.09 (0.59 - 2.04, 0.77) | * |
| 4th quartile | 0.84 (0.43 - 1.64, 0.61) | * |
| State |  |  |
| California | Reference | Reference |
| Connecticut | 0.27 (0.06 - 1.13, 0.07) | * |
| Georgia | 0.64 (0.30 - 1.35, 0.24) | * |
| Hawaii | - | * |
| Iowa | 0.97 (0.42 - 2.25, 0.94) | * |
| Kentucky | 0.63 (0.27 - 1.46, 0.28) | * |
| Louisiana | 1.08 (0.50 - 2.34, 0.84) | * |
| Michigan | 0.24 (0.06 - 1.02, 0.05) | * |
| New Jersey | 0.42 (0.17 - 1.02, 0.06) | * |
| New Mexico | 1.18 (0.28 - 4.99, 0.83) | * |
| Utah | - | * |
| Washington | 0.62 (0.21 - 1.79, 0.38) | * |
| Year of Diagnosis |  |  |
| 2006 | Reference | Reference |
| 2007 | 1.76 (0.78 - 3.98, 0.17) | * |
| 2008 | 1.16 (0.47 - 2.85, 0.75) | * |
| 2009 | 1.81 (0.79 - 4.13, 0.16) | * |
| 2010 | 1.23 (0.49 - 3.10, 0.66) | * |
| 2011 | 1.71 (0.73 - 4.01, 0.21) | * |
| IMRT |  |  |
| No | Reference | Reference |
| Yes | 0.96 (0.55 - 1.68, 0.89) | * |
| ^X^ Multivariate Cox regressions were performed using stepwise forward and backwards elimination with threshold values of p ≤ 0.20 and p ≤ 0.05, respectively.  * Covariate auto-excluded from model during forward or backward selection.  Abbrev: HR, hazard ratio. CI, confidence interval. | | |

| **Supplementary Table T12**. Multivariate proportional hazards regression for cancer-specific survival.^X^ | | |
| --- | --- | --- |
| Parameter | Univariate  HR (95% CI, P-value) | Multivariate  HR (95% CI, P-Value) |
| Daily IGRT |  |  |
| No | Reference | Reference |
| Yes | 0.89 (0.81 - 0.97, 0.01) | 0.94 (0.84 - 1.04, 0.24) |
| Age |  |  |
| 65 - 74 | Reference | Reference |
| 75 - 84 | 1.21 (1.12 - 1.30, <.01) | 1.15 (1.06 - 1.24, <.01) |
| 85+ | 1.52 (1.31 - 1.77, <.01) | 1.26 (1.07 - 1.48, <.01) |
| Race |  |  |
| White | Reference | Reference |
| Black | 0.87 (0.76 - 0.99, 0.04) | * |
| Hispanic | 0.88 (0.58 - 1.34, 0.56) | * |
| Other | 0.92 (0.77 - 1.11, 0.38) | * |
| COPD |  |  |
| No | Reference | Reference |
| Yes | 1.16 (1.08 - 1.25, <.01) | 1.12 (1.03 - 1.21, <.01) |
| Charlson Score (no COPD) |  |  |
| 0 | Reference | Reference |
| 1-2 | 1.03 (0.95 - 1.11, 0.52) | 1.00 (0.93 - 1.09, 0.91) |
| > 2 | 1.27 (1.12 - 1.44, <.01) | 1.19 (1.05 - 1.36, <.01) |
| Supplemental O2 |  |  |
| No | Reference | Reference |
| Yes | 1.23 (1.13 - 1.33, <.01) | 1.10 (1.00 - 1.20, 0.05) |
| Homebound |  |  |
| No | Reference | Reference |
| Yes | 1.19 (0.89 - 1.59, 0.24) | 0.88 (0.65 - 1.18, 0.39) |
| Stage |  |  |
| Stage IIIA | Reference | Reference |
| Stage IIIB | 1.27 (1.18 - 1.36, <.01) | 1.11 (0.98 - 1.25, 0.09) |
| T-Stage |  |  |
| TX | Reference | Reference |
| T0 | 0.37 (0.18 - 0.76, <.01) | 0.43 (0.21 - 0.90, 0.02) |
| T1 | 0.75 (0.62 - 0.91, <.01) | 0.97 (0.77 - 1.21, 0.77) |
| T2 | 0.92 (0.77 - 1.10, 0.34) | 1.04 (0.85 - 1.27, 0.72) |
| T3 | 1.08 (0.88 - 1.32, 0.48) | 1.19 (0.96 - 1.48, 0.12) |
| T4 | 1.14 (0.95 - 1.36, 0.15) | 1.12 (0.90 - 1.39, 0.30) |
| Tumor Size |  |  |
| < 2.0 | Reference | Reference |
| 2.0-5.0 | 1.30 (1.11 - 1.53, <.01) | 1.14 (0.97 - 1.36, 0.12) |
| > 5.0 | 1.62 (1.37 - 1.90, <.01) | 1.40 (1.17 - 1.68, <.01) |
| Unknown | 1.68 (1.41 - 1.99, <.01) | 1.44 (1.18 - 1.76, <.01) |
| Histology |  |  |
| Adenocarcinoma | Reference | Reference |
| SCC | 1.23 (1.13 - 1.34, <.01) | * |
| Large Cell | 1.21 (0.98 - 1.49, 0.08) | * |
| Other | 1.22 (1.11 - 1.35, <.01) | * |
| Laterality |  |  |
| Right | Reference | Reference |
| Left | 0.93 (0.86 - 1.00, 0.05) | 0.91 (0.84 - 0.98, 0.01) |
| Unpaired | 0.93 (0.35 - 2.47, 0.88) | 0.70 (0.26 - 1.89, 0.48) |
| Unknown | 0.60 (0.40 - 0.92, 0.02) | 0.66 (0.42 - 1.05, 0.08) |
| Tumor Location |  |  |
| Main bronchus | Reference | Reference |
| Upper lobe | 0.74 (0.64 - 0.86, <.01) | 0.86 (0.74 - 1.01, 0.06) |
| Middle lobe | 0.89 (0.71 - 1.12, 0.32) | 1.05 (0.83 - 1.32, 0.71) |
| Lower lobe | 0.94 (0.80 - 1.10, 0.41) | 1.06 (0.91 - 1.25, 0.45) |
| Lung NOS | 0.76 (0.62 - 0.94, 0.01) | 0.87 (0.69 - 1.08, 0.21) |
| Other | 1.15 (0.76 - 1.73, 0.51) | 1.33 (0.88 - 2.00, 0.18) |
| PET |  |  |
| No | Reference | Reference |
| Yes | 0.85 (0.74 - 0.97, 0.02) | * |
| # of Positive Nodes |  |  |
| 0 | Reference | Reference |
| 1-3 | 0.99 (0.80 - 1.21, 0.89) | 1.19 (0.96 - 1.47, 0.11) |
| 4+ | 1.06 (0.81 - 1.40, 0.66) | 1.31 (0.99 - 1.74, 0.06) |
| Unknown | 1.56 (1.30 - 1.87, <.01) | 1.43 (1.18 - 1.73, <.01) |
| Treatment Type |  |  |
| Trimodality | Reference | Reference |
| Chemotherapy & radiation | 1.80 (1.54 - 2.09, <.01) | 1.59 (1.32 - 1.91, <.01) |
| Surgery & radiation | 1.44 (1.06 - 1.97, 0.02) | 1.36 (0.99 - 1.86, 0.06) |
| Radiation alone | 2.73 (2.30 - 3.24, <.01) | 2.16 (1.76 - 2.65, <.01) |
| # of RT Fractions |  |  |
| 25 - 29 | Reference | Reference |
| 30 - 34 | 0.92 (0.84 - 1.02, 0.10) | 0.77 (0.70 - 0.85, <.01) |
| 35 - 40 | 0.92 (0.83 - 1.01, 0.09) | 0.76 (0.68 - 0.85, <.01) |
| Type of Treatment Center |  |  |
| Free Standing | Reference | Reference |
| Hospital Based | 0.94 (0.87 - 1.01, 0.11) | * |
| Both | 1.08 (0.69 - 1.70, 0.74) | * |
| Rural vs. Urban |  |  |
| Rural | Reference | Reference |
| Urban | 0.92 (0.84 - 1.00, 0.06) | 0.91 (0.83 - 0.99, 0.04) |
| Radiation Oncologist Density |  |  |
| 1st quartile | Reference | Reference |
| 2nd quartile | 0.99 (0.90 - 1.08, 0.76) | * |
| 3rd quartile | 0.92 (0.83 - 1.02, 0.10) | * |
| 4th quartile | 0.91 (0.81 - 1.01, 0.08) | * |
| Unknown | 0.89 (0.62 - 1.28, 0.54) | * |
| General Surgeon Density |  |  |
| 1st quartile | Reference | Reference |
| 2nd quartile | 1.04 (0.94 - 1.14, 0.47) | * |
| 3rd quartile | 0.98 (0.89 - 1.08, 0.72) | * |
| 4th quartile | 0.96 (0.87 - 1.06, 0.43) | * |
| Unknown | 0.92 (0.64 - 1.33, 0.67) | * |
| Physician Experience |  |  |
| 1st quartile | Reference | Reference |
| 2nd quartile | 1.03 (0.93 - 1.14, 0.53) | * |
| 3rd quartile | 0.99 (0.90 - 1.10, 0.89) | * |
| 4th quartile | 1.06 (0.96 - 1.17, 0.28) | * |
| State |  |  |
| California | Reference | Reference |
| Connecticut | 0.87 (0.74 - 1.03, 0.10) | * |
| Georgia | 1.04 (0.92 - 1.18, 0.52) | * |
| Hawaii | 0.96 (0.66 - 1.38, 0.81) | * |
| Iowa | 1.13 (0.97 - 1.32, 0.13) | * |
| Kentucky | 1.04 (0.91 - 1.19, 0.58) | * |
| Louisiana | 1.20 (1.04 - 1.39, 0.01) | * |
| Michigan | 0.95 (0.81 - 1.10, 0.47) | * |
| New Jersey | 1.11 (0.98 - 1.26, 0.09) | * |
| New Mexico | 0.94 (0.69 - 1.27, 0.67) | * |
| Utah | 0.93 (0.61 - 1.40, 0.72) | * |
| Washington | 0.95 (0.80 - 1.12, 0.55) | * |
| Year of Diagnosis |  |  |
| 2006 | Reference | Reference |
| 2007 | 0.89 (0.80 - 1.00, 0.04) | 0.92 (0.82 - 1.03, 0.15) |
| 2008 | 0.82 (0.73 - 0.92, <.01) | 0.81 (0.72 - 0.91, <.01) |
| 2009 | 0.92 (0.82 - 1.04, 0.18) | 0.94 (0.83 - 1.06, 0.34) |
| 2010 | 0.81 (0.71 - 0.92, <.01) | 0.81 (0.71 - 0.93, <.01) |
| 2011 | 0.61 (0.53 - 0.70, <.01) | 0.61 (0.53 - 0.71, <.01) |
| IMRT |  |  |
| No | Reference | Reference |
| Yes | 1.02 (0.94 - 1.11, 0.65) | 1.13 (1.03 - 1.24, 0.01) |
| ^X^ Multivariate Cox regressions were performed using stepwise forward and backwards elimination with threshold values of p ≤ 0.20 and p ≤ 0.05, respectively.  * Covariate auto-excluded from model during forward or backward selection.  Abbrev: HR, hazard ratio. CI, confidence interval. | | |

| **Supplementary Table T13**. Multivariate proportional hazards regression for overall survival.^X^ | | |
| --- | --- | --- |
| Parameter | Univariate  HR (95% CI, P-value) | Multivariate  HR (95% CI, P-Value) |
| Daily IGRT |  |  |
| No | Reference | Reference |
| Yes | 0.98 (0.90 - 1.06, 0.63) | 0.95 (0.87 - 1.04, 0.29) |
| Age |  |  |
| 65 - 74 | Reference | Reference |
| 75 - 84 | 1.22 (1.15 - 1.31, <.01) | 1.16 (1.08 - 1.24, <.01) |
| 85+ | 1.58 (1.39 - 1.81, <.01) | 1.32 (1.14 - 1.52, <.01) |
| Race |  |  |
| White | Reference | Reference |
| Black | 0.95 (0.85 - 1.07, 0.39) | * |
| Hispanic | 0.96 (0.67 - 1.38, 0.83) | * |
| Other | 0.91 (0.77 - 1.07, 0.25) | * |
| COPD |  |  |
| No | Reference | Reference |
| Yes | 1.19 (1.11 - 1.27, <.01) | 1.12 (1.04 - 1.20, <.01) |
| Charlson Score (no COPD) |  |  |
| 0 | Reference | Reference |
| 1-2 | 1.10 (1.02 - 1.18, <.01) | 1.06 (0.99 - 1.14, 0.11) |
| > 2 | 1.34 (1.20 - 1.50, <.01) | 1.22 (1.09 - 1.37, <.01) |
| Supplemental O2 |  |  |
| No | Reference | Reference |
| Yes | 1.28 (1.19 - 1.38, <.01) | 1.12 (1.04 - 1.22, <.01) |
| Homebound |  |  |
| No | Reference | Reference |
| Yes | 1.39 (1.09 - 1.77, <.01) | 1.04 (0.81 - 1.33, 0.76) |
| Stage |  |  |
| Stage IIIA | Reference | Reference |
| Stage IIIB | 1.20 (1.13 - 1.28, <.01) | 1.06 (0.95 - 1.18, 0.29) |
| T-Stage |  |  |
| TX | Reference | Reference |
| T0 | 0.51 (0.30 - 0.88, 0.02) | 0.52 (0.30 - 0.90, 0.02) |
| T1 | 0.80 (0.67 - 0.96, 0.01) | 0.99 (0.81 - 1.21, 0.94) |
| T2 | 0.96 (0.82 - 1.13, 0.65) | 1.07 (0.89 - 1.28, 0.48) |
| T3 | 1.05 (0.88 - 1.27, 0.58) | 1.16 (0.95 - 1.41, 0.15) |
| T4 | 1.13 (0.96 - 1.33, 0.14) | 1.14 (0.94 - 1.38, 0.19) |
| Tumor Size |  |  |
| < 2.0 | Reference | Reference |
| 2.0-5.0 | 1.24 (1.08 - 1.42, <.01) | 1.09 (0.94 - 1.26, 0.26) |
| > 5.0 | 1.48 (1.29 - 1.71, <.01) | 1.30 (1.11 - 1.52, <.01) |
| Unknown | 1.54 (1.32 - 1.79, <.01) | 1.35 (1.13 - 1.60, <.01) |
| Histology |  |  |
| Adenocarcinoma | Reference | Reference |
| SCC | 1.23 (1.14 - 1.32, <.01) | * |
| Large Cell | 1.09 (0.90 - 1.33, 0.36) | * |
| Other | 1.19 (1.09 - 1.31, <.01) | * |
| Laterality |  |  |
| Right | Reference | Reference |
| Left | 0.95 (0.89 - 1.02, 0.15) | * |
| Unpaired | 0.76 (0.29 - 2.03, 0.59) | * |
| Unknown | 0.70 (0.49 - 0.99, 0.04) | * |
| Tumor Location |  |  |
| Main bronchus | Reference | Reference |
| Upper lobe | 0.78 (0.68 - 0.89, <.01) | 0.91 (0.79 - 1.04, 0.16) |
| Middle lobe | 0.85 (0.69 - 1.05, 0.14) | 1.04 (0.84 - 1.29, 0.72) |
| Lower lobe | 0.95 (0.82 - 1.09, 0.46) | 1.08 (0.93 - 1.25, 0.32) |
| Lung NOS | 0.77 (0.64 - 0.93, <.01) | 0.84 (0.69 - 1.02, 0.08) |
| Other | 1.16 (0.80 - 1.68, 0.44) | 1.37 (0.94 - 2.00, 0.10) |
| PET |  |  |
| No | Reference | Reference |
| Yes | 0.84 (0.74 - 0.94, <.01) | * |
| # of Positive Nodes |  |  |
| 0 | Reference | Reference |
| 1-3 | 1.02 (0.85 - 1.23, 0.79) | 1.17 (0.97 - 1.42, 0.10) |
| 4+ | 1.07 (0.84 - 1.37, 0.59) | 1.26 (0.98 - 1.62, 0.07) |
| Unknown | 1.58 (1.35 - 1.86, <.01) | 1.41 (1.19 - 1.68, <.01) |
| Treatment Type |  |  |
| Trimodality | Reference | Reference |
| Chemotherapy & radiation | 1.72 (1.50 - 1.96, <.01) | 1.54 (1.31 - 1.80, <.01) |
| Surgery & radiation | 1.37 (1.05 - 1.81, 0.02) | 1.33 (1.00 - 1.75, 0.05) |
| Radiation alone | 2.61 (2.25 - 3.03, <.01) | 2.08 (1.74 - 2.48, <.01) |
| # of RT Fractions |  |  |
| 25 - 29 | Reference | Reference |
| 30 - 34 | 0.93 (0.85 - 1.01, 0.09) | 0.77 (0.70 - 0.84, <.01) |
| 35 - 40 | 0.92 (0.84 - 1.01, 0.08) | 0.76 (0.69 - 0.84, <.01) |
| Type of Treatment Center |  |  |
| Free Standing | Reference | Reference |
| Hospital Based | 0.96 (0.90 - 1.03, 0.24) | * |
| Both | 1.18 (0.80 - 1.76, 0.41) | * |
| Rural vs. Urban |  |  |
| Rural | Reference | Reference |
| Urban | 0.91 (0.84 - 0.98, 0.02) | 0.90 (0.83 - 0.98, 0.02) |
| Radiation Oncologist Density |  |  |
| 1st quartile | Reference | Reference |
| 2nd quartile | 0.98 (0.90 - 1.07, 0.65) | * |
| 3rd quartile | 0.93 (0.85 - 1.01, 0.09) | * |
| 4th quartile | 0.91 (0.83 - 1.00, 0.06) | * |
| Unknown | 0.93 (0.67 - 1.27, 0.64) | * |
| General Surgeon Density |  |  |
| 1st quartile | Reference | Reference |
| 2nd quartile | 1.04 (0.95 - 1.13, 0.39) | * |
| 3rd quartile | 0.98 (0.90 - 1.07, 0.67) | * |
| 4th quartile | 0.96 (0.87 - 1.05, 0.35) | * |
| Unknown | 0.96 (0.70 - 1.32, 0.80) | * |
| Physician Experience |  |  |
| 1st quartile | Reference | Reference |
| 2nd quartile | 1.02 (0.94 - 1.12, 0.60) | * |
| 3rd quartile | 1.02 (0.93 - 1.12, 0.69) | * |
| 4th quartile | 1.00 (0.91 - 1.09, 0.99) | * |
| State |  |  |
| California | Reference | Reference |
| Connecticut | 0.91 (0.79 - 1.05, 0.18) | * |
| Georgia | 1.06 (0.95 - 1.18, 0.32) | * |
| Hawaii | 1.00 (0.72 - 1.38, 0.99) | * |
| Iowa | 1.11 (0.96 - 1.28, 0.16) | * |
| Kentucky | 1.03 (0.92 - 1.16, 0.60) | * |
| Louisiana | 1.22 (1.07 - 1.40, <.01) | * |
| Michigan | 0.98 (0.86 - 1.12, 0.80) | * |
| New Jersey | 1.09 (0.98 - 1.22, 0.12) | * |
| New Mexico | 1.09 (0.85 - 1.41, 0.49) | * |
| Utah | 0.94 (0.65 - 1.36, 0.75) | * |
| Washington | 0.94 (0.81 - 1.09, 0.39) | * |
| Year of Diagnosis |  |  |
| 2006 | Reference | Reference |
| 2007 | 0.93 (0.84 - 1.03, 0.15) | 0.96 (0.86 - 1.06, 0.39) |
| 2008 | 0.85 (0.76 - 0.94, <.01) | 0.84 (0.75 - 0.93, <.01) |
| 2009 | 0.99 (0.89 - 1.10, 0.85) | 1.00 (0.90 - 1.12, 0.95) |
| 2010 | 0.91 (0.82 - 1.03, 0.13) | 0.92 (0.81 - 1.03, 0.16) |
| 2011 | 0.90 (0.80 - 1.01, 0.07) | 0.90 (0.79 - 1.02, 0.09) |
| IMRT |  |  |
| No | Reference | Reference |
| Yes | 1.07 (0.99 - 1.15, 0.10) | 1.12 (1.03 - 1.22, 0.01) |
| ^X^ Multivariate Cox regressions were performed using stepwise forward and backwards elimination with threshold values of p ≤ 0.20 and p ≤ 0.05, respectively.  * Covariate auto-excluded from model during forward or backward selection.  Abbrev: HR, hazard ratio. CI, confidence interval. | | |

| **Supplementary Table T14**. Bivariate distribution table of 6-month survivor cohort used in cost analysis. | | | | | | | | | | |
| --- | --- | --- | --- | --- | --- | --- | --- | --- | --- | --- |
| Characteristic | **Unmatched** | | | | | **Matched** | | | | |
|  | IGRT (N) | IGRT (%) | No IGRT (N) | No IGRT (%) | P-value | IGRT (N) | IGRT (%) | No IGRT (N) | No IGRT (%) | P-value |
| Age |  |  |  |  | 0.56 |  |  |  |  | 0.95 |
| 65 - 74 | 1697 | 56.6 | 467 | 54.8 |  | 328 | 53.4 | 324 | 52.8 |  |
| 75 - 84 | 1135 | 37.9 | 340 | 39.9 |  | 255 | 41.5 | 257 | 41.9 |  |
| 85+ | 165 | 5.5 | 45 | 5.3 |  | 31 | 5 | 33 | 5.4 |  |
| Sex |  |  |  |  | 0.84 |  |  |  |  | 0.05 |
| Male | 1596 | 53.3 | 457 | 53.6 |  | 304 | 49.5 | 339 | 55.2 |  |
| Female | 1401 | 46.7 | 395 | 46.4 |  | 310 | 50.5 | 275 | 44.8 |  |
| Race |  |  |  |  | 0.04 |  |  |  |  | 0.06 |
| White | 2574 | 85.9 | ≥743 | ≥87.2 |  | ≥536 | ≥87.3 | 557 | 90.7 |  |
| Black | 267 | 8.9 | 68 | 8 |  | 52 | 8.5 | 42 | 6.8 |  |
| Hispanic | 29 | 1 | ≤11 | ≤1.3 |  | ≤11 | ≤1.8 | 0 | 0 |  |
| Other | 127 | 4.2 | 30 | 3.5 |  | 15 | 2.4 | 15 | 2.4 |  |
| State |  |  |  |  | <.01 |  |  |  |  | 0.14 |
| California | 680 | 22.7 | ≥155 | ≥18.2 |  | ≥113 | ≥18.4 | ≥71 | ≥11.5 |  |
| Connecticut | 224 | 7.5 | 51 | 6 |  | 42 | 6.8 | 50 | 8.1 |  |
| Georgia | 444 | 14.8 | 124 | 14.6 |  | 84 | 13.7 | 70 | 11.4 |  |
| Hawaii | 39 | 1.3 | ≤11 | ≤1.3 |  | ≤11 | ≤1.8 | ≤11 | ≤1.8 |  |
| Iowa | 195 | 6.5 | 64 | 7.5 |  | 52 | 8.5 | 61 | 9.9 |  |
| Kentucky | 312 | 10.4 | 86 | 10.1 |  | 48 | 7.8 | 68 | 11.1 |  |
| Louisiana | 230 | 7.7 | 63 | 7.4 |  | 49 | 8 | 53 | 8.6 |  |
| Michigan | 228 | 7.6 | 74 | 8.7 |  | 58 | 9.4 | 70 | 11.4 |  |
| New Jersey | 393 | 13.1 | 128 | 15 |  | 80 | 13 | 81 | 13.2 |  |
| New Mexico | 53 | 1.8 | ≤11 | ≤1.3 |  | 12 | 2 | ≤11 | ≤1.8 |  |
| Utah | 26 | 0.9 | ≤11 | ≤1.3 |  | ≤11 | ≤1.8 | ≤11 | ≤1.8 |  |
| Washington | 173 | 5.8 | 74 | 8.7 |  | 54 | 8.8 | 57 | 9.3 |  |
| Local Coverage Determination |  |  |  |  | 0.92 |  |  |  |  | 0.28 |
| Favorable | 653 | 21.8 | 190 | 22.3 |  | 140 | 22.8 | 134 | 21.8 |  |
| Intermediate | 1773 | 59.2 | 504 | 59.2 |  | 366 | 59.6 | 350 | 57 |  |
| Unfavorable | 571 | 19.1 | 158 | 18.5 |  | 108 | 17.6 | 130 | 21.2 |  |
| Year of Diagnosis |  |  |  |  | <.01 |  |  |  |  | 0.92 |
| 2006 | 661 | 22.1 | 12 | 1.4 |  | ≤11 | ≤1.8 | 12 | 2 |  |
| 2007 | 630 | 21 | 66 | 7.7 |  | 55 | 9 | 66 | 10.7 |  |
| 2008 | 575 | 19.2 | 94 | 11 |  | 97 | 15.8 | 93 | 15.1 |  |
| 2009 | 468 | 15.6 | 179 | 21 |  | 136 | 22.1 | 135 | 22 |  |
| 2010 | 358 | 11.9 | 204 | 23.9 |  | 158 | 25.7 | 151 | 24.6 |  |
| 2011 | 305 | 10.2 | 297 | 34.9 |  | ≥157 | ≥25.5 | 157 | 25.6 |  |
| Charlson Score (No COPD) |  |  |  |  | 0.06 |  |  |  |  | 0.35 |
| 0 | 1714 | 57.2 | 449 | 52.7 |  | 348 | 56.7 | 323 | 52.6 |  |
| 1-2 | 1022 | 34.1 | 319 | 37.4 |  | 210 | 34.2 | 232 | 37.8 |  |
| > 2 | 261 | 8.7 | 84 | 9.9 |  | 56 | 9.1 | 59 | 9.6 |  |
| COPD |  |  |  |  | 0.36 |  |  |  |  | 0.39 |
| No | 1653 | 55.2 | 455 | 53.4 |  | 352 | 57.3 | 337 | 54.9 |  |
| Yes | 1344 | 44.8 | 397 | 46.6 |  | 262 | 42.7 | 277 | 45.1 |  |
| Supplemental O2 |  |  |  |  | 0.22 |  |  |  |  | 0.41 |
| No | 2275 | 75.9 | 664 | 77.9 |  | 472 | 76.9 | 484 | 78.8 |  |
| Yes | 722 | 24.1 | 188 | 22.1 |  | 142 | 23.1 | 130 | 21.2 |  |
| Homebound | 50 | 1.7 | ≤11 | ≤1.3 | <.01 | ≤11 | ≤1.8 | ≤11 | ≤1.8 | 1.00 |
| Histology |  |  |  |  | <.01 |  |  |  |  | 0.89 |
| Adenocarcinoma | 982 | 32.8 | 340 | 39.9 |  | 225 | 36.6 | 237 | 38.6 |  |
| SCC | 1236 | 41.2 | 341 | 40 |  | 257 | 41.9 | 251 | 40.9 |  |
| Large Cell | 98 | 3.3 | 20 | 2.3 |  | 16 | 2.6 | 17 | 2.8 |  |
| Other | 681 | 22.7 | 151 | 17.7 |  | 116 | 18.9 | 109 | 17.8 |  |
| Stage |  |  |  |  | 0.32 |  |  |  |  | 0.19 |
| Stage IIIA | 1508 | 50.3 | 445 | 52.2 |  | 297 | 48.4 | 320 | 52.1 |  |
| Stage IIIB | 1489 | 49.7 | 407 | 47.8 |  | 317 | 51.6 | 294 | 47.9 |  |
| T-Stage |  |  |  |  | 0.68 |  |  |  |  | 0.44 |
| TX | 127 | 4.2 | 41 | 4.8 |  | 21 | 3.4 | 31 | 5 |  |
| T0 | 17 | 0.6 | ≤11 | ≤1.3 |  | ≤11 | ≤1.8 | ≤11 | ≤1.8 |  |
| T1 | 435 | 14.5 | 123 | 14.4 |  | 95 | 15.5 | 92 | 15 |  |
| T2 | 991 | 33.1 | 300 | 35.2 |  | 190 | 30.9 | 204 | 32.7 |  |
| T3 | 288 | 9.6 | 82 | 9.6 |  | 56 | 9.1 | 64 | 10.4 |  |
| T4 | 1139 | 38 | ≥295 | ≥34.6 |  | ≥241 | ≥39.3 | ≥212 | ≥34.5 |  |
| Tumor Size |  |  |  |  | 0.62 |  |  |  |  | 0.63 |
| < 2.0 | 226 | 7.5 | 58 | 6.8 |  | 40 | 6.5 | 36 | 5.9 |  |
| 2.0-5.0 | 1399 | 46.7 | 393 | 46.1 |  | 279 | 45.4 | 293 | 47.7 |  |
| > 5.0 | 939 | 31.3 | 285 | 33.5 |  | 218 | 35.5 | 200 | 32.6 |  |
| Unknown | 433 | 14.4 | 116 | 13.6 |  | 77 | 12.5 | 85 | 13.8 |  |
| Tumor Laterality |  |  |  |  | 0.87 |  |  |  |  | 0.94 |
| Right | ≥1726 | ≥57.6 | ≥494 | ≥58 |  | ≥335 | ≥55.3 | ≥345 | ≥56.2 |  |
| Left | 1228 | 41 | 337 | 39.6 |  | 257 | 41.9 | 247 | 40.2 |  |
| Unpaired | ≤11 | ≤0.4 | ≤11 | ≤1.3 |  | ≤11 | ≤1.8 | ≤11 | ≤1.8 |  |
| Unknown | 32 | 1.1 | 10 | 1.2 |  | ≤11 | ≤1.8 | ≤11 | ≤1.8 |  |
| Tumor Location |  |  |  |  | 0.04 |  |  |  |  | 0.22 |
| Main bronchus | 178 | 5.9 | 42 | 4.9 |  | 38 | 6.2 | 27 | 4.4 |  |
| Upper lobe | 1766 | 58.9 | ≥525 | ≥61.6 |  | ≥358 | ≥58.3 | ≥369 | ≥60.1 |  |
| Middle lobe | 119 | 4 | 23 | 2.7 |  | 18 | 2.9 | 16 | 2.6 |  |
| Lower lobe | 752 | 25.1 | 204 | 23.9 |  | 161 | 26.2 | 155 | 25.2 |  |
| Lung NOS | 155 | 5.2 | 47 | 5.5 |  | 28 | 4.6 | 36 | 5.9 |  |
| Other | 27 | 0.9 | ≤11 | ≤1.3 |  | ≤11 | ≤1.8 | ≤11 | ≤1.8 |  |
| PET | 2778 | 92.7 | 801 | 94 | 0.18 | 575 | 93.6 | 574 | 93.5 | 0.91 |
| # of nodes positive |  |  |  |  | 0.50 |  |  |  |  | 0.85 |
| 0 | 146 | 4.9 | 37 | 4.3 |  | 28 | 4.6 | 27 | 4.4 |  |
| 1-3 | 391 | 13 | 118 | 13.8 |  | 70 | 11.4 | 77 | 12.5 |  |
| 4+ | 104 | 3.5 | 22 | 2.6 |  | 20 | 3.3 | 16 | 2.6 |  |
| Unknown | 2356 | 78.6 | 675 | 79.2 |  | 496 | 80.8 | 494 | 80.5 |  |
| Treatment Type |  |  |  |  | 0.19 |  |  |  |  | 0.37 |
| Trimodality | 269 | 9 | 67 | 7.9 |  | 62 | 10.1 | 47 | 7.7 |  |
| Chemotherapy & radiation | 2216 | 73.9 | 661 | 77.6 |  | ≥457 | ≥74.4 | ≥472 | ≥76.9 |  |
| Surgery & radiation | 58 | 1.9 | 15 | 1.8 |  | ≤11 | ≤1.8 | ≤11 | ≤1.8 |  |
| Radiation alone | 454 | 15.1 | 109 | 12.8 |  | 84 | 13.7 | 89 | 14.5 |  |
| # of RT Fractions |  |  |  |  | <.01 |  |  |  |  | 0.83 |
| 25 - 29 | 540 | 18 | 162 | 19 |  | 124 | 20.2 | 118 | 19.2 |  |
| 30 - 34 | 1348 | 45 | 316 | 37.1 |  | 248 | 40.4 | 244 | 39.7 |  |
| 35 - 40 | 1109 | 37 | 374 | 43.9 |  | 242 | 39.4 | 252 | 41 |  |
| IMRT | 422 | 14.1 | 476 | 55.9 | <.01 | 263 | 42.8 | 266 | 43.3 | 0.86 |
| Type of Treatment Center |  |  |  |  | <.01 |  |  |  |  | 0.80 |
| Free-Standing | 948 | 31.6 | 355 | 41.7 |  | 226 | 36.9 | 220 | 35.8 |  |
| Hospital Based | 2028 | 67.7 | ≥486 | ≥57 |  | ≥377 | ≥61.4 | ≥383 | ≥62.3 |  |
| Both | 21 | 0.7 | ≤11 | ≤1.3 |  | ≤11 | ≤1.8 | ≤11 | ≤1.8 |  |
| Rural vs. Urban |  |  |  |  | 0.08 |  |  |  |  | 0.27 |
| Rural | 574 | 19.2 | 141 | 16.5 |  | 109 | 17.8 | 124 | 20.2 |  |
| Urban | 2423 | 80.8 | 711 | 83.5 |  | 505 | 82.2 | 490 | 79.8 |  |
| Radiation Oncologist Density |  |  |  |  | <.01 |  |  |  |  | 0.19 |
| 1st quartile | 961 | 32.1 | 257 | 30.2 |  | ≥191 | ≥31.2 | ≥175 | ≥28.5 |  |
| 2nd quartile | 772 | 25.8 | ≥254 | ≥29.8 |  | 184 | 30 | 160 | 26.1 |  |
| 3rd quartile | 699 | 23.3 | 197 | 23.1 |  | 144 | 23.5 | 173 | 28.2 |  |
| 4th quartile | 526 | 17.6 | 133 | 15.6 |  | 84 | 13.7 | 95 | 15.5 |  |
| Unknown | 39 | 1.3 | ≤11 | ≤1.3 |  | ≤11 | ≤1.8 | ≤11 | ≤1.8 |  |
| General Surgeon Density |  |  |  |  | <.01 |  |  |  |  | 0.78 |
| 1st quartile | 950 | 31.7 | 219 | 25.7 |  | 175 | 28.5 | 168 | 27.4 |  |
| 2nd quartile | 700 | 23.4 | 204 | 23.4 |  | 134 | 21.8 | 134 | 21.8 |  |
| 3rd quartile | 703 | 23.5 | ≥226 | ≥27 |  | ≥177 | ≥28.9 | ≥168 | ≥27.3 |  |
| 4th quartile | 605 | 20.2 | 192 | 22.5 |  | 117 | 19.1 | 133 | 21.7 |  |
| Unknown | 39 | 1.3 | ≤11 | ≤1.3 |  | ≤11 | ≤1.8 | ≤11 | ≤1.8 |  |
| Physician Experience |  |  |  |  | <.01 |  |  |  |  | 0.20 |
| 1st quartile | ≥803 | ≥26.8 | 155 | 18.2 |  | 135 | 22 | 122 | 19.9 |  |
| 2nd quartile | 756 | 25.2 | 210 | 24.6 |  | 185 | 30.1 | 162 | 26.4 |  |
| 3rd quartile | 668 | 22.3 | 304 | 35.7 |  | 159 | 25.9 | 186 | 30.3 |  |
| 4th quartile | 759 | 25.3 | 183 | 21.5 |  | 135 | 22 | 144 | 23.5 |  |
| Other | ≤11 | ≤0.4 | 0 | 0 |  | 0 | 0 | 0 | 0 |  |
| * To protect patient anonymity, all cells with values between 1 and 11 were masked with the indicator “≤11.” Then the largest cell in the same column for the same characteristic was adjusted in the opposite direction so that the total number of observations in that column remained the same. | | | | | | | | | | |

| **Supplementary Table T15**. Costs of treatment | | | | | | | |
| --- | --- | --- | --- | --- | --- | --- | --- |
|  |  | **Cost ($)** | | | | | |
|  |  | *hfIGRT* | *No hfIGRT* | *Difference* | *Lower 95% CI* | *Upper 95% CI* | *P-value* |
| **All Patients** | *IGRT* | $2,248.60 | $197.17 | $2,051.43 | $1,916.46 | $2,179.23 | < 0.01 |
|  | *Radiation* | $19,083.44 | $12,009.1 | $7,074.34 | $6,559.47 | $7,581.49 | < 0.01 |
|  | *All Care* | $74,587.75 | $64,839.15 | $9,748.6 | $6,237.24 | $13,426.86 | < 0.01 |
| **Matched Cohort** | *IGRT* | $1,971.41 | $199.03 | $1,772.38 | $1,605.69 | $1,925.71 | < 0.01 |
|  | *Radiation* | $17,330.31 | $15,024.17 | $2,306.15 | $1,559.12 | $3,046.65 | < 0.01 |
|  | *All Care* | $71,568.63 | $69,693.25 | $1,875.38 | -$3,230.72 | $6,924.93 | 0.49 |

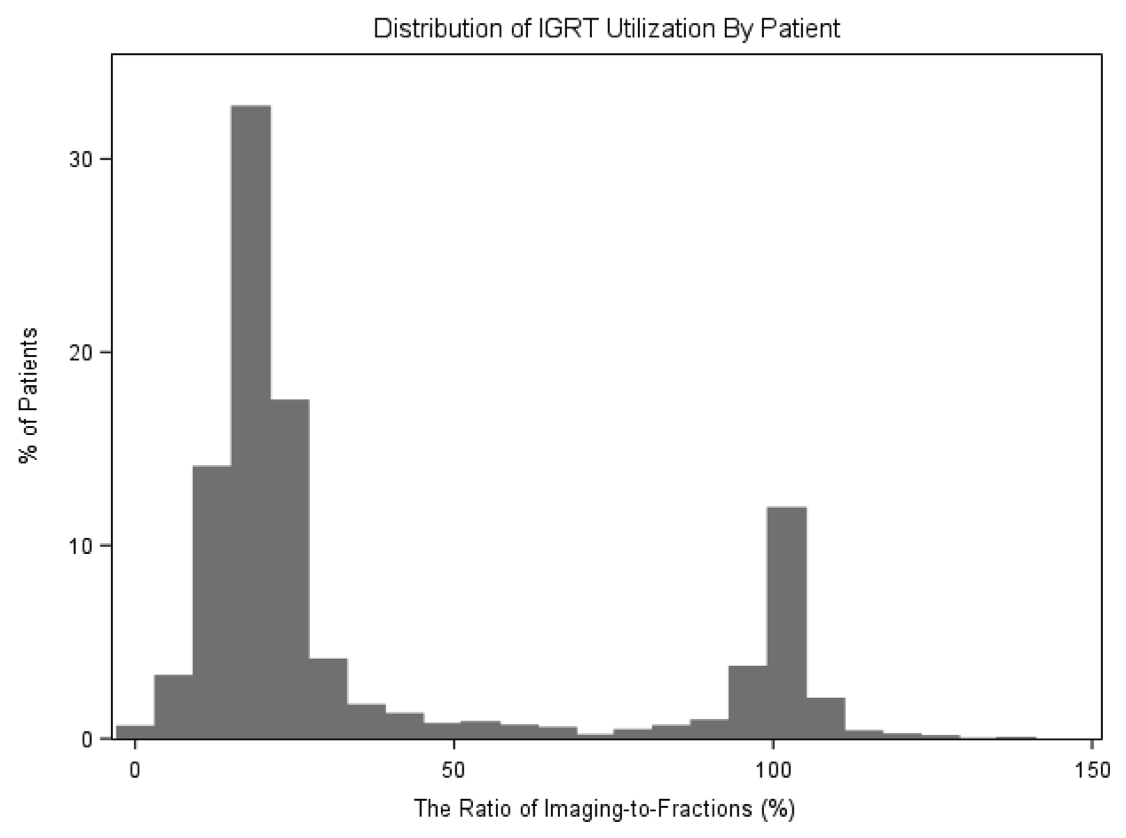
**Supplementary** **Figure F1**. Distribution of IGRT Utilization by Patient

**Figure F1**: Histogram showing the % of patients using IGRT at different frequencies. X-axis represents the percentage of radiation fractions accompanied by image-guidance. Y-axis is the percent of patients in our cohort who received IGRT at that frequency. The distribution is bimodal, with most patient receiving image guidance with ~20% of radiation fractions or ~100% of radiation fractions. Some patients appear to receive image guidance with greater than 100% of radiation fractions because initial planning CT scans are often billed using the same code as IGRT and cannot be distinguished using billing, even though these CT scans are not accompanied by a radiation fraction.

**Supplementary** **Figure F2**. Billing for Port/KV vs. CBCT Films for IGRT Over Time

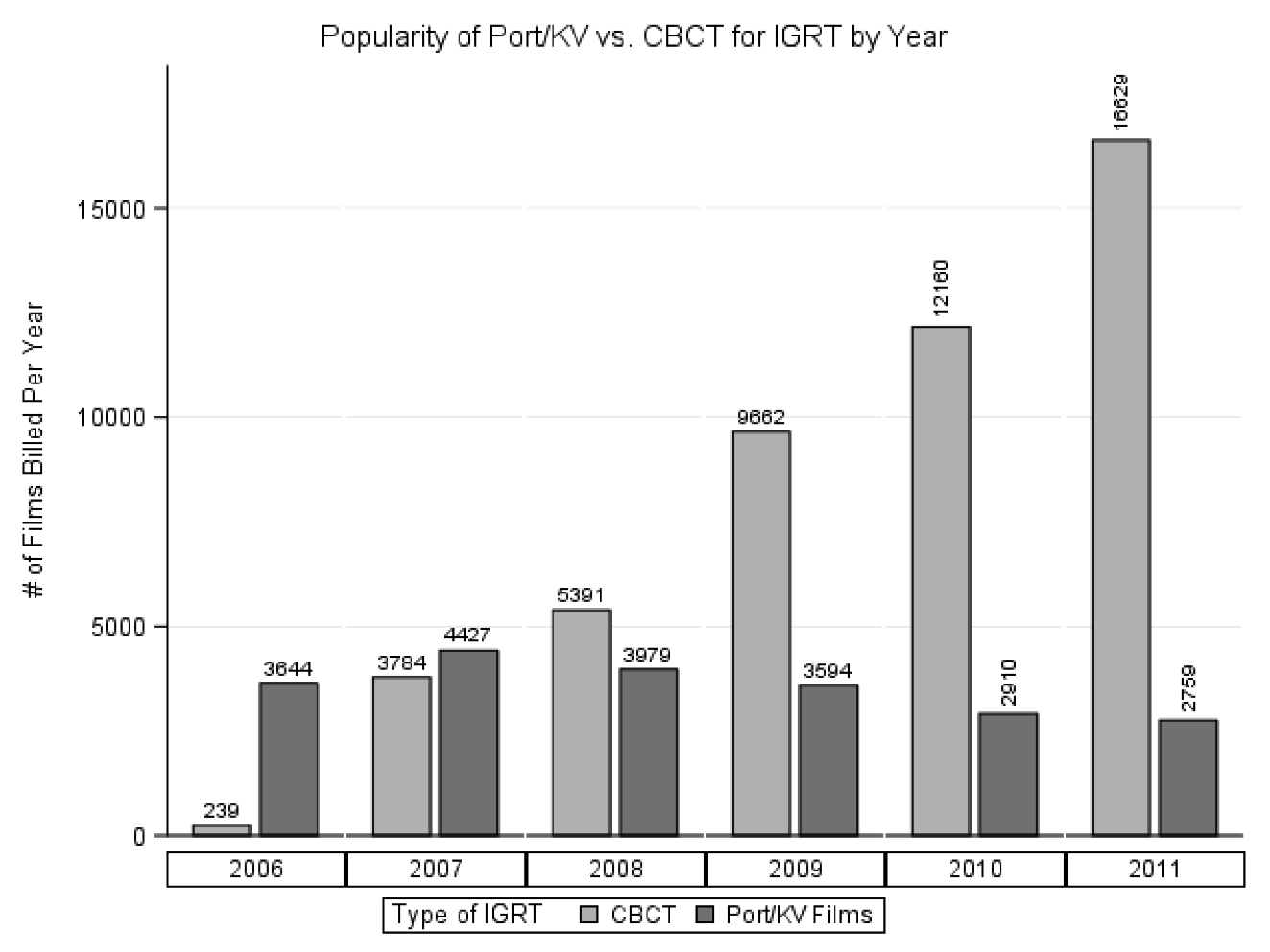
**Figure F2**: Total number of Medicare claims for CBCT and Port/KV based image guidance in our cohort over time. CBCT rapidly overtook Port/KV imaging as the preferred method of image guidance.

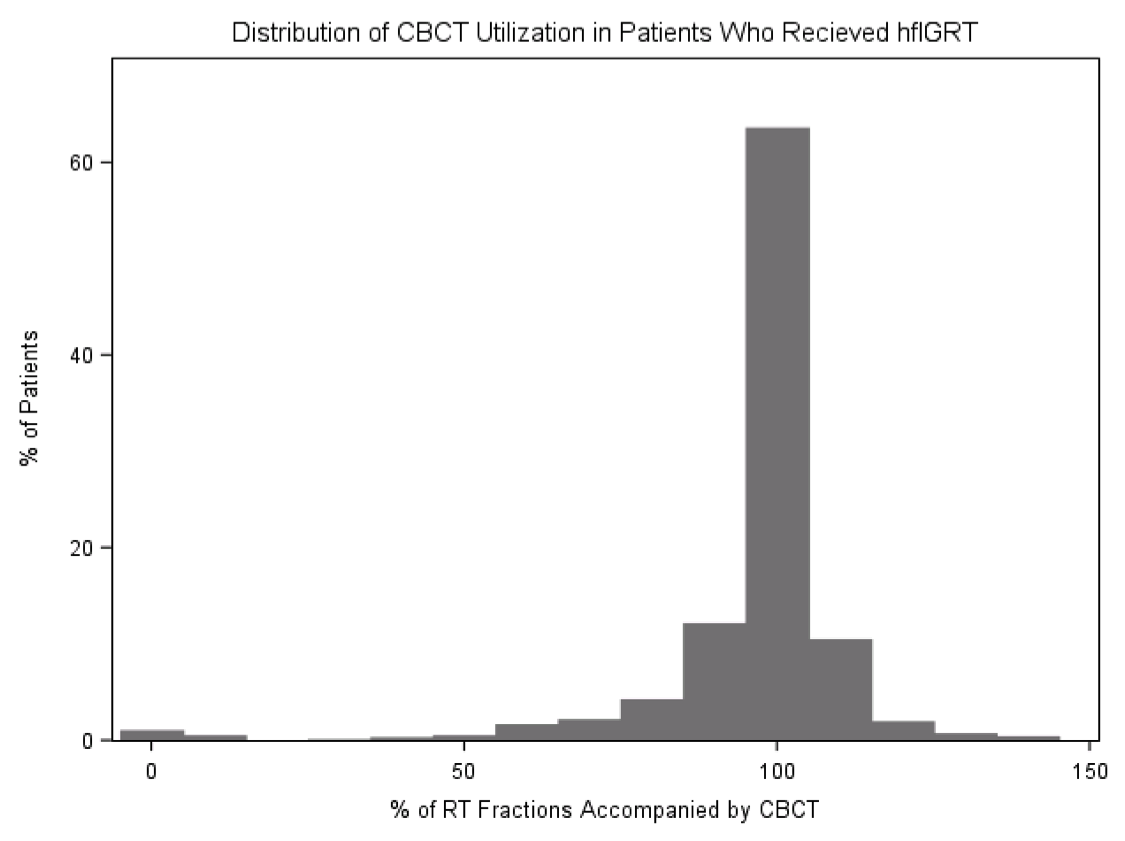
**Supplementary** **Figure F3**. Distribution of CBCT Utilization in Patients Who Received hfIGRT.

**Figure F3**: % of radiation fractions accompanied by CBCT in patient who are receiving hfIGRT. Clearly the vast majority of patients who are receiving hfIGRT are getting most of their fractions with CBCT imaging.

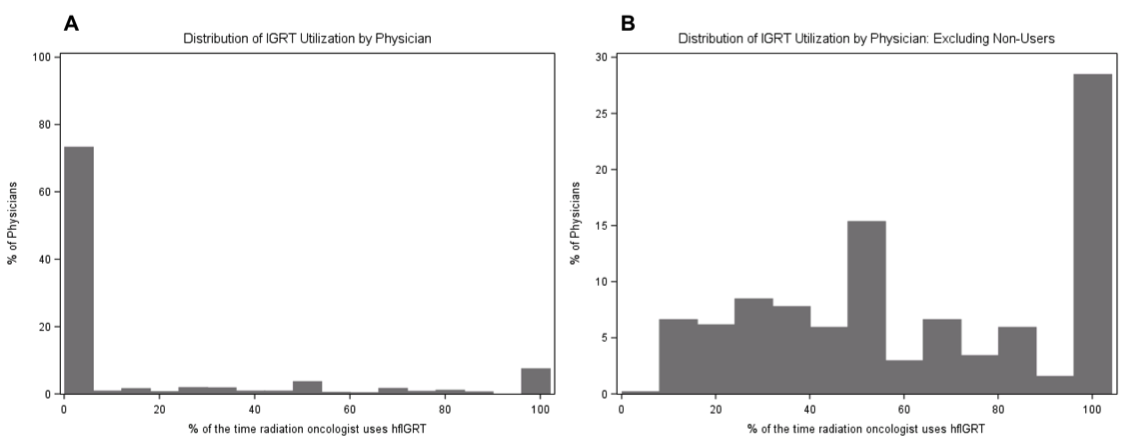
**Supplementary** **Figure F4**. Distribution of IGRT Utilization by Physician

**Figure F4**: (A) Histogram showing the distribution of IGRT utilization by physicians. The x-axis shows the percentage of stage III NSCLC patients each radiation oncologist uses hfIGRT in. The Y axis represent the percentage of radiation oncologist who use hfIGRT with that frequency. (B) The same as figure 4A, except excluding radiation oncologist who used hfIGRT in none of their patients.

**Supplementary** **Figure F5**. IGRT Cost by LCD

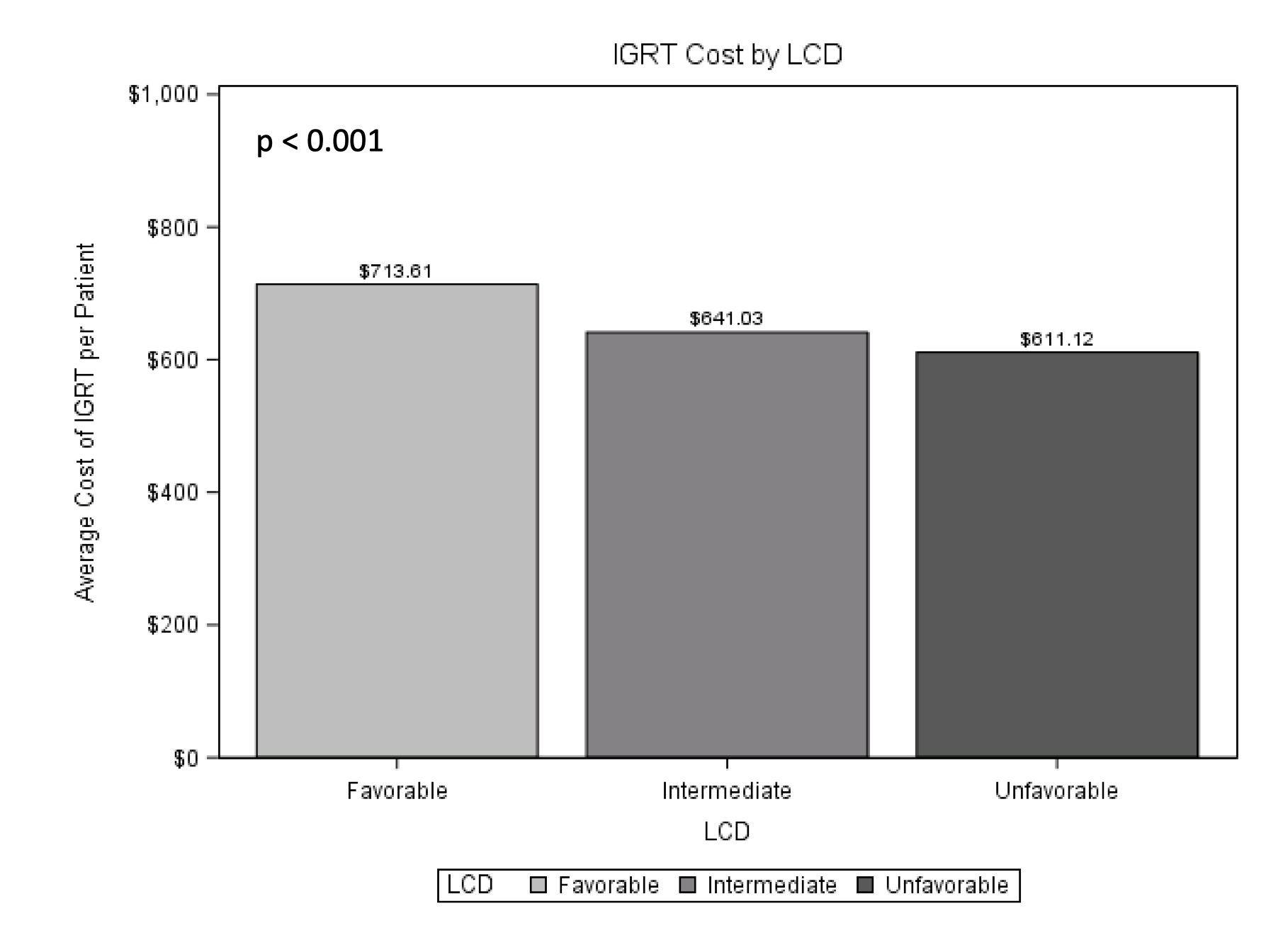

**Figure F5**: Shows the average cost of image guidance in IMRT favorable, intermediate, and unfavorable regions of the united states. Costs were calculated for each patient by summing the Medicare reimbursement for all image guidance associated bills.

**REFRENCES FOR THE APPENDIX**

1.	<https://healthcaredelivery.cancer.gov/seermedicare/program/comorbidity.html>. NCIDoCCPSS-MCoCWa.

2.	Charlson ME, Sax FL, MacKenzie CR, Fields SD, Braham RL, Douglas RG, Jr. Assessing illness severity: does clinical judgment work? *J Chronic Dis.* 1986;39(6):439-452.

3.	Klabunde CN, Legler JM, Warren JL, Baldwin LM, Schrag D. A refined comorbidity measurement algorithm for claims-based studies of breast, prostate, colorectal, and lung cancer patients. *Ann Epidemiol.* 2007;17(8):584-590.

4.	Klabunde CN, Potosky AL, Legler JM, Warren JL. Development of a comorbidity index using physician claims data. *J Clin Epidemiol.* 2000;53(12):1258-1267.

5.	Centers for Medicare & Medicaid Services (CMS) Chronic Conditions Data Warehouse (CCW) Chronic Condition Algorithms. Definitions at <https://www.ccwdata.org/web/guest/condition-categories>.

6.	Warren JL, Harlan LC, Fahey A, et al. Utility of the SEER-Medicare data to identify chemotherapy use. *Med Care.* 2002;40(8 Suppl):IV-55-61.

7.	Boero IJ, Paravati AJ, Xu B, et al. Importance of radiation oncologist experience among patients with head-and-neck cancer treated with intensity-modulated radiation therapy. *Journal of Clinical Oncology.* 2016;34(7):684-690.
